# Physical and mental health disability associated with long-COVID: Baseline results from a US nationwide cohort

**DOI:** 10.1101/2022.12.07.22283203

**Authors:** Bryan Lau, Eryka Wentz, Zhanmo Ni, Karine Yenokyan, Candelaria Coggiano, Shruti H. Mehta, Priya Duggal

**Affiliations:** Department of Epidemiology, Johns Hopkins Bloomberg School of Public Health, Baltimore, MD

## Abstract

**Importance:** Persistent symptoms after SARS-COV-2 infection, or long-COVID, may occur in anywhere from 10-55% of those who have had COVID-19, but the extent of impact on daily functioning and disability remains unquantified.

**Objective:** To characterize physical and mental disability associated with long-COVID

**Design:** Cross-sectional analysis of baseline data from a cohort study

**Setting:** Online US nationwide survey

**Participants:** Adults 18 years of age and older who live in the US who either report a history of COVID-19 illness (n=8,874) or report never having had COVID-19 (n=633)

**Main Outcome and Measures:** Self-reported mobility disability (difficulty walking a quarter of a mile and/or up 10 stairs, instrumental activities of daily living [IADL] disability (difficulty doing light or heavy housework), and mental fatigue as measured by the Wood Mental Fatigue Inventory (WMFI).

**Results:** Of 7,926 participants with long-COVID, the median age was 45 years, 84% were female, 89% self-reported white race, and 7.4% self-reported Hispanic/Latino ethnicity. Sixty-five percent of long-COVID participants were classified as having at least one disability, compared to 6% of those with resolved-COVID (n=948) and 14% of those with no-COVID (n=633). Of long-COVID participants, about 1% and 5% were classified as critically physically disabled or mentally fatigued, respectively. Age, prior comorbidity, increased BMI, female gender, hospitalization for COVID-19, non-white race, and multi-race were all associated with significantly higher disability burden. Dizziness at the time of infection (33% non-hospitalized, 39% hospitalized) was associated with all five disability components in both hospitalized and non-hospitalized groups. Heavy limbs, dyspnea, and tremors were associated with four of the five components of disability in the non-hospitalized group, and heavy limbs was associated with four of the five components in the hospitalized group. Vaccination was protective against development of disability.

**Conclusion and Relevance:** We observed a high burden of physical and mental disability associated with long-COVID which has serious implications for individual and societal health that may be partially mitigated by vaccination. Longitudinal characterization and evaluation of COVID-19 patients is necessary to identify patterns of recovery and treatment options.

## Introduction

Much of the public health response to COVID-19 has been focused on the acute phase of infection, from curtailing transmission to preventing and treating severe disease. However, research suggests that post-acute sequelae after SARS-CoV-2 infection, or long-COVID, can occur in anywhere from 10 to 55%^1–3^ and has the potential to overwhelm health systems and economies. While data consistently suggest that long-COVID disproportionately affects those hospitalized at the time of their infection, it is increasingly recognized that persistent symptoms affect those with mild/moderate infections. Moreover, long-COVID also affects all ages, sexes, and races and occurs among individuals without pre-existing comorbidity.^4–6^ Symptoms reported by individuals with long-COVID are heterogeneous and affect multiple organ systems.^7,8^

While reports have characterized the breadth of symptoms that persist or newly occur in long-COVID, there have been fewer assessments on downstream impacts such as disability. In one study of critically ill hospitalized COVID-19 patients, 48% had decreased functional status and 10% reported severe limitations in their daily life six months post-infection^9^. Similarly, in a Norwegian cohort of hospitalized older patients, 35% reported impaired ability to perform daily activities, 33% reduced mobility, and 43% a decrease in cognitive function six months after hospitalization.^10^ While there have been fewer assessments from community-based settings, one study of 328 participants reported that 4.9% had symptoms post-infection that constrained daily activities^11^. This functional restriction can directly affect employment, caregiving, and independent living.

We characterize physical disability and mental fatigue in those with and without COVID-19 in a large sample of community dwelling adults living in the US from the Johns Hopkins COVID Long Study.

## METHODS

### Study design and sample

The Johns Hopkins COVID Long Study is an online prospective cohort of adults. The study launched on February 2, 2021 and expanded to include those without a history of COVID-19 on March 11, 2022. Participants must be at least 18 years of age and reside in the US. For COVID-19 focused questions, participants must also self-report a positive SARS-CoV-2 test or symptoms of COVID-19. The study was approved by the Johns Hopkins Bloomberg School of Public Health Institutional Review Board and all participants provide informed consent.

Recruitment was done primarily through social media posts (e.g., Instagram, Facebook, and Twitter) and Facebook ad campaigns. Additionally, we used direct messaging through emails to health departments, religious institutions, and via word of mouth. We also enrolled the study in the Johns Hopkins Opportunities for Participant Engagement (HOPE) Registry which matches individuals interested in participating in COVID-19 research with open studies (http://johnshopkinshope.org/).

Between February 2, 2021, and May 27, 2022, 18,666 participants with a history of COVID-19 consented; 897 participants without a history of COVID-19 consented between March 11, 2022, and July 8, 2022. For this analysis, we used baseline data from participants with long-COVID (n=7,926), resolved-COVID (n=948), and without a history of COVID-19 or no-COVID (n=633) who had complete data on the three disability outcomes (mobility disability, instrumental activities of daily living (IADL) disability, and mental fatigue) and did not report disability prior to infection (Figure S1.1). Compared to those who had complete data, those missing data were no different with respect to age, gender, race/ethnicity, region, education, employment, occupation, or income. To define long-COVID, we used the World Health Organization (WHO) definition^12^ of having at least one symptom 12 weeks post initial infection.

### Data collection

The baseline survey collected data on socio-demographics, COVID-19 testing and symptoms, physician-diagnosed conditions post-infection, COVID-19 treatments and hospitalizations, comorbidities, physical limitations, vaccines, sleep quality, anxiety, and mental fatigue. All data were collected in REDCap.^13,14^

### Case Definitions

For mobility and IADL disability, we used the classification by the Baltimore Longitudinal Study of Aging.^15^ For mobility disability, participants were asked about difficulty walking ¼ mile and/or climbing 10 stairs, and if they reported difficulty, the level (a little, some, a lot, unable to do). Mobility disability was defined as having *some or greater level of difficulty* walking ¼ mile and/or climbing 10 stairs. For IADL disability, participants were asked about difficulty with light housework and heavy housework, and if so, the level of difficulty (a little, some, a lot, unable to do). IADL disability was defined as having at least *some or greater level of difficulty* with heavy housework. Critical physical disability was defined as being unable to do mobility or IADL tasks.

To assess mental fatigue, we utilized the Wood Mental Fatigue Inventory (WMFI)^16^ which included nine domains. Participants were assigned a score for each domain: 0 (not at all), 1 (a little), 2 (somewhat), 3 (quite a lot), and 4 (very much). The scores were summed with higher scores representing worse mental fatigue with a cutoff of 20 based on the 90th percentile of the no-COVID sample (Supplement Methods).

### Statistical Analysis

We used inverse odds weights to ensure that the distribution of variables within the resolved-COVID and no-COVID sample matched our long-COVID sample^17^. We focused on three outcomes - incident mobility, IADL and mental fatigue disability. Logistic regression was used to examine the association of demographics, prior comorbidities, BMI, and prior activity level with each outcome in individual models among those with long-COVID. We further evaluated presenting symptoms from the acute phase of infection with each disability outcome. Any questions added to the baseline survey after individuals enrolled were accounted for by single stochastic imputation, since missingness was completely random. As the list of presenting symptoms was large and comprehensive, we used three methods for symptom variable selection: 1) variable importance using a random forest with 95% confidence intervals to select variables that were influential in classifying disability^18^ from a multivariate outcome random forest, 2) a multinomial model with relaxed least absolute shrinkage and selection operator (LASSO), and 3) a sequential logistic model with relaxed LASSO.

Finally, we investigated the effect of vaccination status at the time of infection on the development of long-COVID disability. We weighted the analysis to be representative of the US population through the COVID-19 Symptom Survey (Supplement Methods).^19^ Two approaches were used: a regression-based model informed by directed acyclic graphs (DAGs) and an instrumental variable (IV). Vaccination was defined into three categories (unvaccinated/one mRNA dose, two mRNA doses/one adenoviral dose, or additional vaccine doses (i.e., boosters)). Due to sample size in the IV analyses, we used a binary category (≥2 mRNA doses or ≥1 dose of adenoviral vector vaccine vs. unvaccinated/one mRNA dose).

## RESULTS

Among 7,926 individuals with long-COVID, the median age was 45 years, 84% were female, 89% self-reported white race, and 7.4% self-reported Hispanic/Latino ethnicity (Table 1). Pre-existing comorbidities were heterogeneous with participants reporting hypertension (15%), diabetes (4.0%), and asthma/other chronic pulmonary conditions (17%). Based on calculated body mass index, 36% were classified as obese. Comparable characteristics of persons with resolved-COVID and no-COVID are included in Table 1.

**Table 1.**
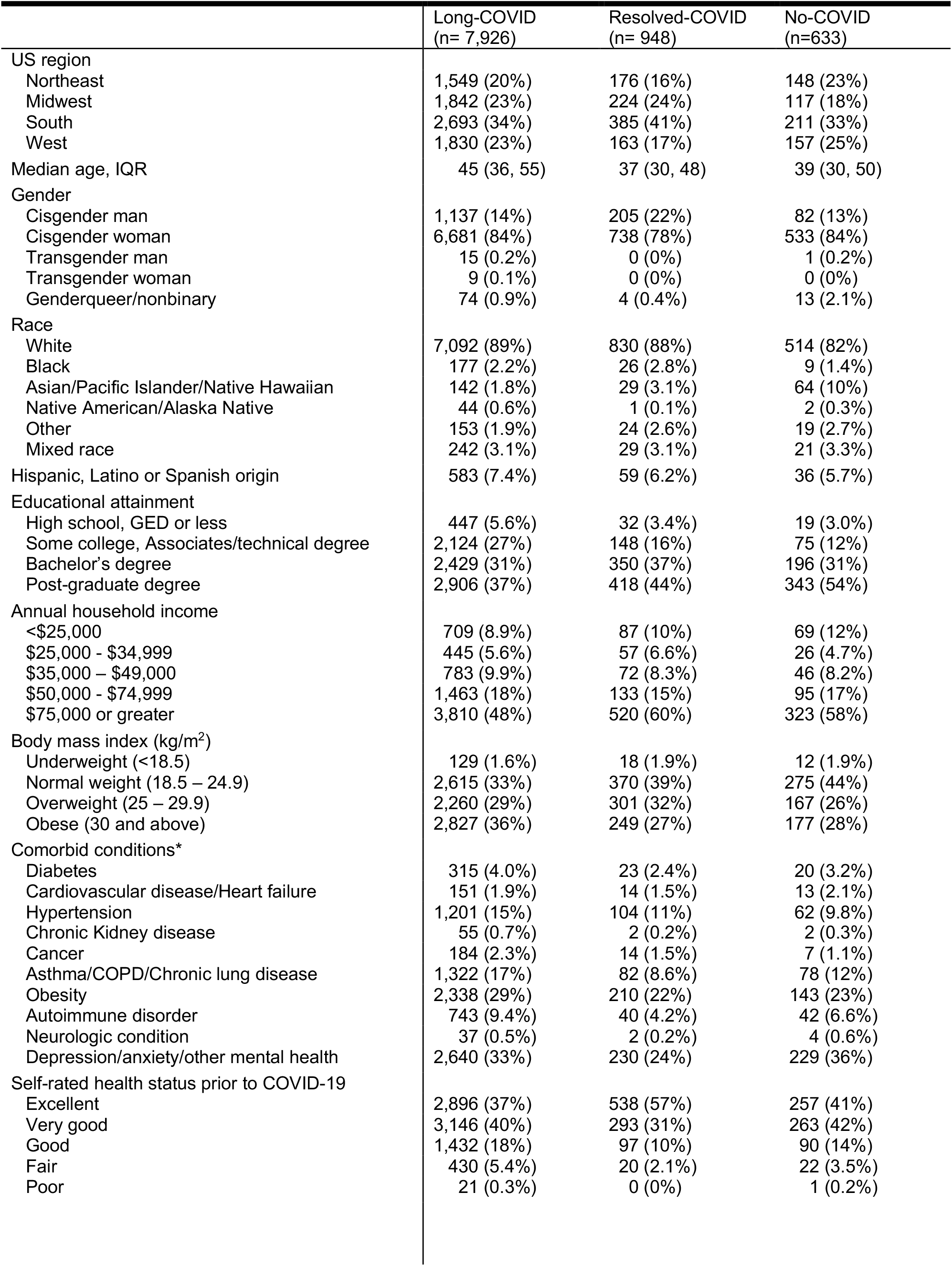
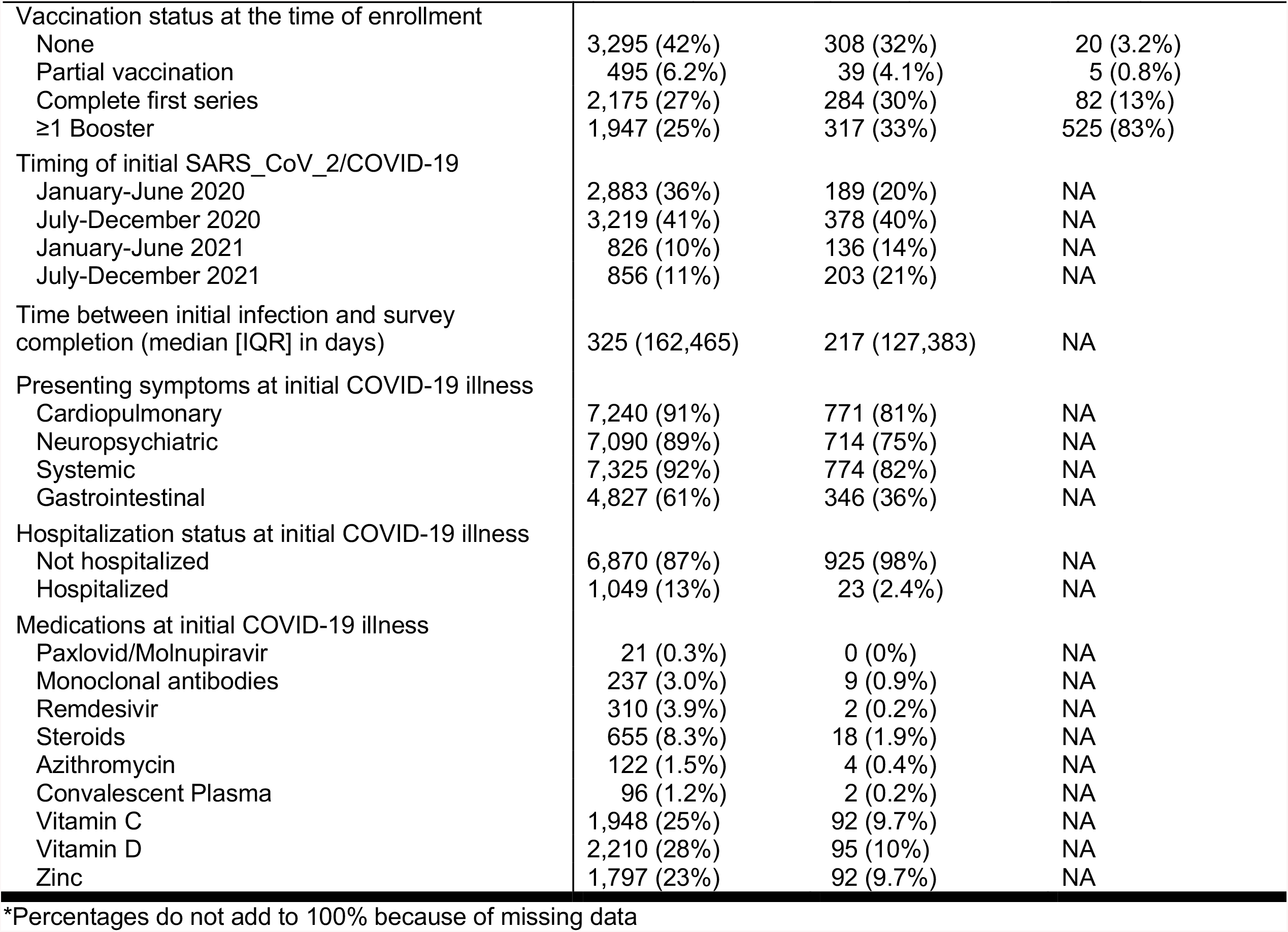
Characteristics of participants in the JHU COVID Long Study*

|  | Long-COVID<br>(n= 7,926) | Resolved-COVID<br>(n= 948) | No-COVID<br>(n=633) |
| --- | --- | --- | --- |
| US region |  |  |  |
| Northeast | 1,549 (20%) | 176 (16%) | 148 (23%) |
| Midwest | 1,842 (23%) | 224 (24%) | 117 (18%) |
| South | 2,693 (34%) | 385 (41%) | 211 (33%) |
| West | 1,830 (23%) | 163 (17%) | 157 (25%) |
| Median age, IQR | 45 (36, 55) | 37 (30, 48) | 39 (30, 50) |
| Gender |  |  |  |
| Cisgender man | 1,137 (14%) | 205 (22%) | 82 (13%) |
| Cisgender woman | 6,681 (84%) | 738 (78%) | 533 (84%) |
| Transgender man | 15 (0.2%) | 0 (0%) | 1 (0.2%) |
| Transgender woman | 9 (0.1%) | 0 (0%) | 0 (0%) |
| Genderqueer/nonbinary | 74 (0.9%) | 4 (0.4%) | 13 (2.1%) |
| Race |  |  |  |
| White | 7,092 (89%) | 830 (88%) | 514 (82%) |
| Black | 177 (2.2%) | 26 (2.8%) | 9 (1.4%) |
| Asian/Pacific Islander/Native Hawaiian | 142 (1.8%) | 29 (3.1%) | 64 (10%) |
| Native American/Alaska Native | 44 (0.6%) | 1 (0.1%) | 2 (0.3%) |
| Other | 153 (1.9%) | 24 (2.6%) | 19 (2.7%) |
| Mixed race | 242 (3.1%) | 29 (3.1%) | 21 (3.3%) |
| Hispanic, Latino or Spanish origin | 583 (7.4%) | 59 (6.2%) | 36 (5.7%) |
| Educational attainment |  |  |  |
| High school, GED or less | 447 (5.6%) | 32 (3.4%) | 19 (3.0%) |
| Some college, Associates/technical degree | 2,124 (27%) | 148 (16%) | 75 (12%) |
| Bachelor's degree | 2,429 (31%) | 350 (37%) | 196 (31%) |
| Post-graduate degree | 2,906 (37%) | 418 (44%) | 343 (54%) |
| Annual household income |  |  |  |
| <\$25,000 | 709 (8.9%) | 87 (10%) | 69 (12%) |
| \$25,000 - \$34,999 | 445 (5.6%) | 57 (6.6%) | 26 (4.7%) |
| \$35,000 – \$49,000 | 783 (9.9%) | 72 (8.3%) | 46 (8.2%) |
| \$50,000 - \$74,999 | 1,463 (18%) | 133 (15%) | 95 (17%) |
| \$75,000 or greater | 3,810 (48%) | 520 (60%) | 323 (58%) |
| Body mass index (kg/m<sup>2</sup>) |  |  |  |
| Underweight (<18.5) | 129 (1.6%) | 18 (1.9%) | 12 (1.9%) |
| Normal weight (18.5 – 24.9) | 2,615 (33%) | 370 (39%) | 275 (44%) |
| Overweight (25 – 29.9) | 2,260 (29%) | 301 (32%) | 167 (26%) |
| Obese (30 and above) | 2,827 (36%) | 249 (27%) | 177 (28%) |
| Comorbid conditions* |  |  |  |
| Diabetes | 315 (4.0%) | 23 (2.4%) | 20 (3.2%) |
| Cardiovascular disease/Heart failure | 151 (1.9%) | 14 (1.5%) | 13 (2.1%) |
| Hypertension | 1,201 (15%) | 104 (11%) | 62 (9.8%) |
| Chronic Kidney disease | 55 (0.7%) | 2 (0.2%) | 2 (0.3%) |
| Cancer | 184 (2.3%) | 14 (1.5%) | 7 (1.1%) |
| Asthma/COPD/Chronic lung disease | 1,322 (17%) | 82 (8.6%) | 78 (12%) |
| Obesity | 2,338 (29%) | 210 (22%) | 143 (23%) |
| Autoimmune disorder | 743 (9.4%) | 40 (4.2%) | 42 (6.6%) |
| Neurologic condition | 37 (0.5%) | 2 (0.2%) | 4 (0.6%) |
| Depression/anxiety/other mental health | 2,640 (33%) | 230 (24%) | 229 (36%) |
| Self-rated health status prior to COVID-19 |  |  |  |
| Excellent | 2,896 (37%) | 538 (57%) | 257 (41%) |
| Very good | 3,146 (40%) | 293 (31%) | 263 (42%) |
| Good | 1,432 (18%) | 97 (10%) | 90 (14%) |
| Fair | 430 (5.4%) | 20 (2.1%) | 22 (3.5%) |
| Poor | 21 (0.3%) | 0 (0%) | 1 (0.2%) |
| Vaccination status at the time of enrollment |  |  |  |
| None | 3,295 (42%) | 308 (32%) | 20 (3.2%) |
| Partial vaccination | 495 (6.2%) | 39 (4.1%) | 5 (0.8%) |
| Complete first series | 2,175 (27%) | 284 (30%) | 82 (13%) |
| ≥1 Booster | 1,947 (25%) | 317 (33%) | 525 (83%) |
| Timing of initial SARS_CoV_2/COVID-19 |  |  |  |
| January-June 2020 | 2,883 (36%) | 189 (20%) | NA |
| July-December 2020 | 3,219 (41%) | 378 (40%) | NA |
| January-June 2021 | 826 (10%) | 136 (14%) | NA |
| July-December 2021 | 856 (11%) | 203 (21%) | NA |
| Time between initial infection and survey completion (median [IQR] in days) | 325 (162,465) | 217 (127,383) | NA |
| Presenting symptoms at initial COVID-19 illness |  |  |  |
| Cardiopulmonary | 7,240 (91%) | 771 (81%) | NA |
| Neuropsychiatric | 7,090 (89%) | 714 (75%) | NA |
| Systemic | 7,325 (92%) | 774 (82%) | NA |
| Gastrointestinal | 4,827 (61%) | 346 (36%) | NA |
| Hospitalization status at initial COVID-19 illness |  |  |  |
| Not hospitalized | 6,870 (87%) | 925 (98%) | NA |
| Hospitalized | 1,049 (13%) | 23 (2.4%) | NA |
| Medications at initial COVID-19 illness |  |  |  |
| Paxlovid/Molnupiravir | 21 (0.3%) | 0 (0%) | NA |
| Monoclonal antibodies | 237 (3.0%) | 9 (0.9%) | NA |
| Remdesivir | 310 (3.9%) | 2 (0.2%) | NA |
| Steroids | 655 (8.3%) | 18 (1.9%) | NA |
| Azithromycin | 122 (1.5%) | 4 (0.4%) | NA |
| Convalescent Plasma | 96 (1.2%) | 2 (0.2%) | NA |
| Vitamin C | 1,948 (25%) | 92 (9.7%) | NA |
| Vitamin D | 2,210 (28%) | 95 (10%) | NA |
| Zinc | 1,797 (23%) | 92 (9.7%) | NA |

\*Percentages do not add to 100% because of missing data

Initial infection spanned from January 2020-June 2022, with the majority (77%) occurring before January 2021 and vaccine availability. At initial COVID-19 illness, 89% of those with long-COVID and 73% of those with resolved-COVID had not been vaccinated. However, at enrollment, 58% of those with long-COVID and 67% of those with resolved-COVID had completed at least one (adenovirus) or two (mRNA) vaccine doses. The five most reported acute infection symptoms for those with long-COVID were lack of energy (86%), headache (71%), muscle aches (61%), fever (54%), and shortness of breath/dyspnea (52%). While the types of symptoms were comparable for those with resolved-COVID, only 2.4% with resolved-COVID required hospitalization compared to 13% with long-COVID (Table 1).

Prior to COVID-19, 77% of long-COVID participants reported that their general health status was ‘very good’ or ‘excellent’, yet only 6.6% of individuals in this group reported the same health status as before infection, compared to 81% of those with resolved-COVID (Figure 1A). Most individuals reported being physically active before infection (71% of long-COVID, 66% of resolved-COVID). However, 50% of individuals with long-COVID experienced declines in their physical activity, including 33% moving from ‘active’ to ‘sedentary’ compared to 3% among those with resolved-COVID (Figure 1B).

**Figure 1.**
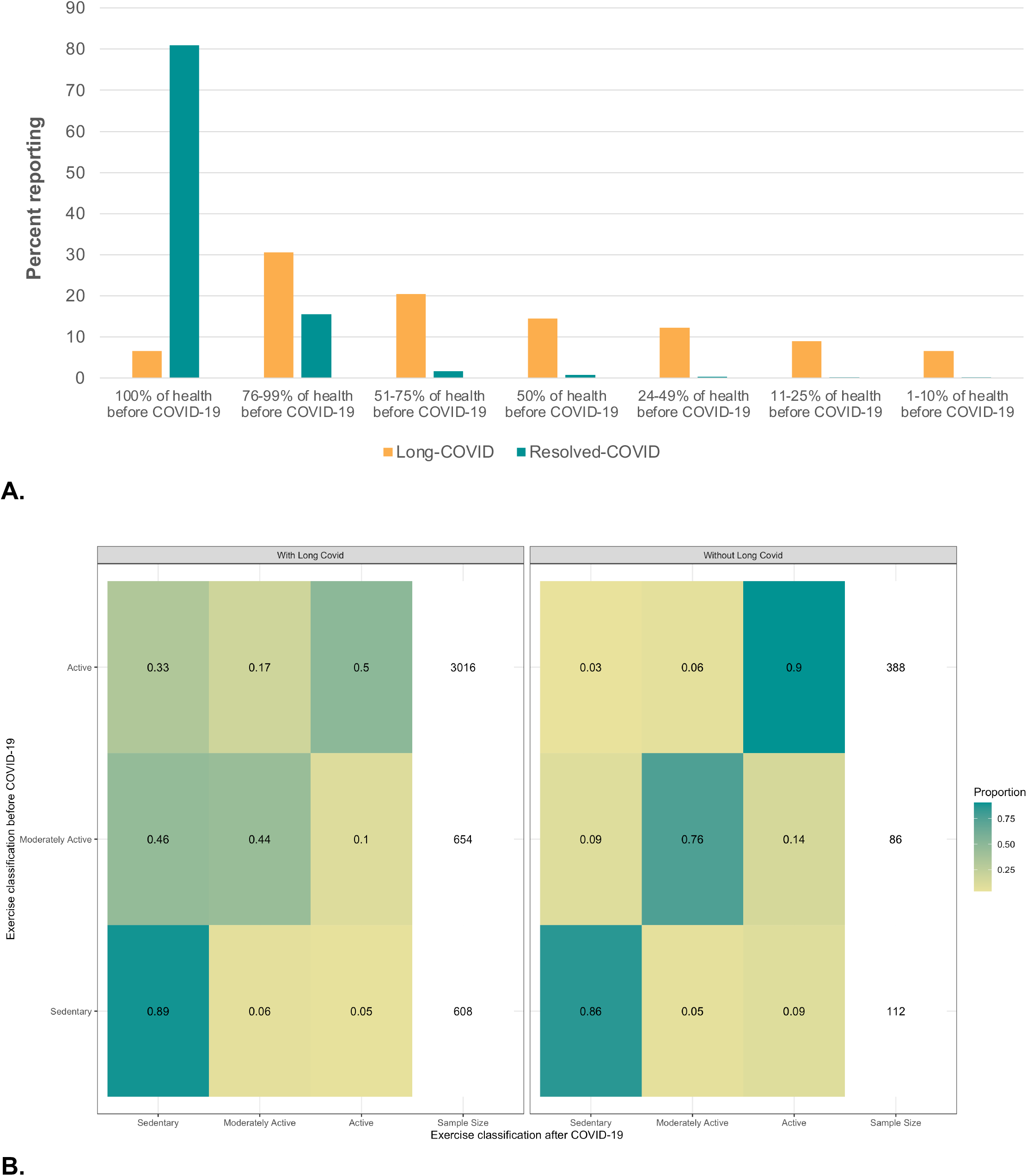
Self-reported changes in health status and physical activity since COVID-19 illness among persons with long-COVID and resolved-COVID. A: Self-reported health relative to pre-COVID health among persons with long-covid vs. resolved-covid. B: Self-reported exercise level before and after COVID-19 illness among persons with long covid vs. resolved-covid.

### COVID-19 status and disability

Of long-COVID participants, 65% were classified as having at least one disability compared to 6% of those with resolved-COVID and 14% of those with no-COVID. Those requiring hospitalization (n=1049) had higher prevalence of disability (68% mobility, 80% IADL, 47% mental fatigue) but the prevalence was also elevated in those not hospitalized (n=6870, 37% mobility, 53% IADL, 33% mental fatigue). Of those with at least one incident disability (Figure S7), 22% were categorized with all three disabilities (hospitalized 38%, non-hospitalized 19%).

Of those with long-COVID, 41% were classified as mobility disabled, compared to 2.7% of resolved-COVID and 4.5% of no-COVID (Figure 2). In the long-COVID group, 6.1% indicated they could not walk ¼ mile or up 10 stairs, and 0.9% were incapable of doing both. For IADL, 57% of those with long-COVID had at least some IADL difficulty with heavy and/or light housework compared to 2.7% and 10% for resolved-COVID and no-COVID, respectively (Figure 2). Of those with long-COVID, 12% were unable to do either light or heavy housework and 1.4% could not complete either. The median WMFI score was 15 (IQR: 7-24), 2 (IQR: 0-6) and 5 (IQR: 2-12) for long-COVID, resolved-COVID, and no-COVID, respectively (Figure 2). Of those with long-COVID, 6.8% were classified with severe mental fatigue (WMFI 30-34) and 4.7% critically severe (WMFI 35-36) compared to 0.2%(severe) and 0% (critically severe) for those with resolved COVID and 0.9% (severe) and 0.3% (critically severe) for those with no-COVID. Across the spectrum of severity, participants reported new physician diagnosis since infection with the highest burden among those classified as critically disabled. The most reported new diagnosis was tachycardia, followed by postural-orthostatic tachycardia syndrome, myalgic encephalomyelitis/chronic fatigue syndrome, and irritable bowel syndrome (Figure S10.1-S10.3).

**Figure 2.**
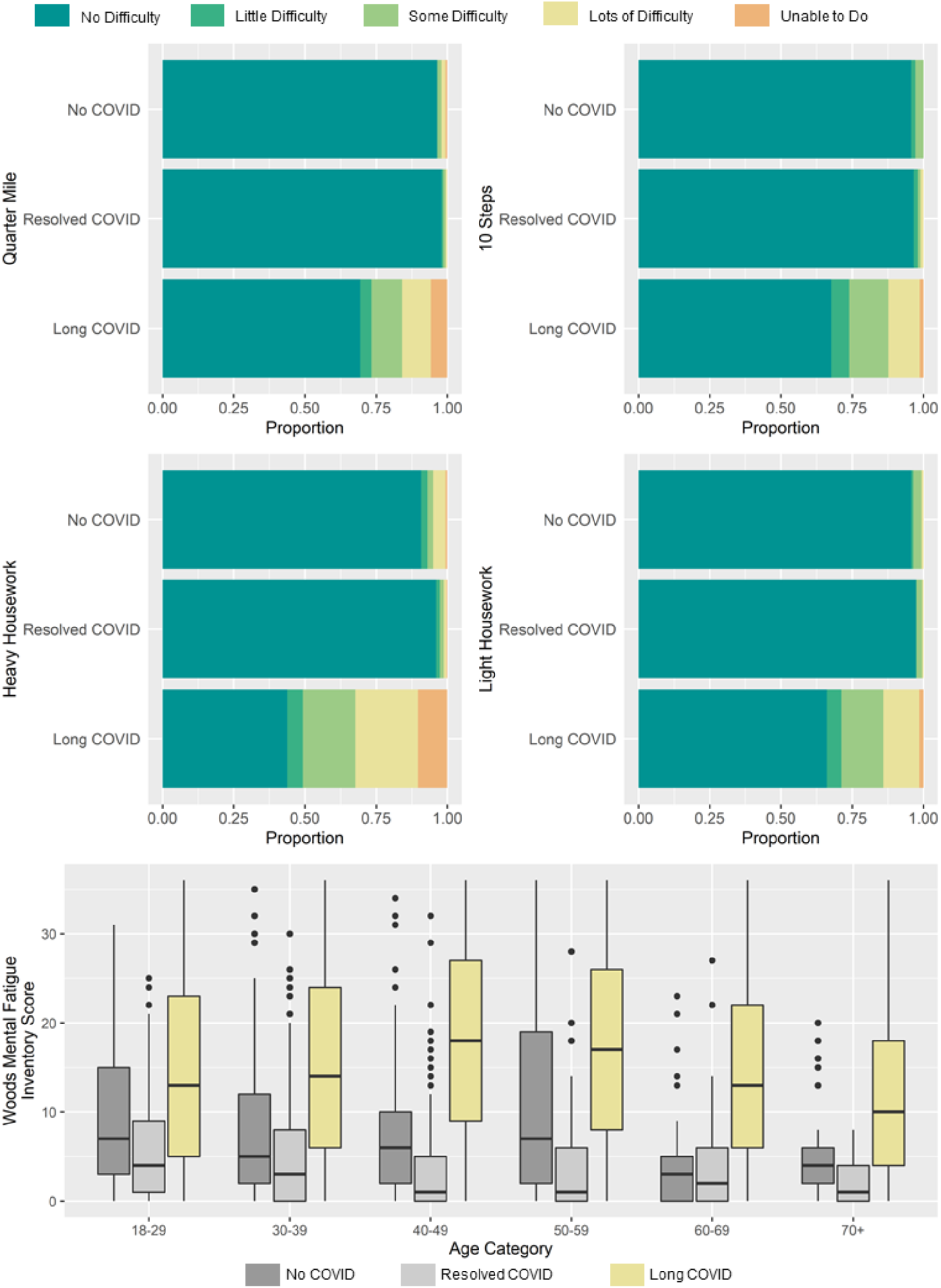
Disability measures by COVID-19 status: long-COVID, resolved-COVID, and no-COVID. A: Level of difficulty in walking a quarter of a mile. B: Level of difficulty in walking up ten stairs; C: Level of difficulty in doing heavy housework. D: Level of difficulty in doing light housework. E: Level of mental fatigue (WMFI score) by age. All estimates are weighted in order to standardize covariates of the group without long-COVID and the group with no history of COVID-19 to that of the long-COVID sample. Factors that were standardized included: age, gender, BMI, whether individuals were hospitalized during their acute phase of infection, and prior co-morbidity, specifically diabetes, cardiovascular disease, history of heart attack, congestive heart failure, high blood pressure, high cholesterol, history of stroke, autoimmune disorders, Hepatitis C, asthma, chronic lung disease, chronic kidney disease, cancer, depression, being pregnant, overweight or obese, anxiety or other mental health condition, and chronic or acute Lyme disease.

Several non-symptom factors increased odds of all three disabilities including age, prior comorbidity, increased BMI, female gender, COVID-19 hospitalization, education, and income. When we stratified mobility disability by hospitalization status, only BMI, female gender, some levels of education, and some levels of income were associated among hospitalized participants. Similarly, when we stratified IADL disability and mental fatigue by hospitalization status, only female gender, some levels of education, and some levels of income was associated for IADL and age for mental fatigue. (Table S8.1-S8.3).

### Symptoms during Acute Infection and Disability

We further examined associations between symptoms reported during initial infection with each of the components of the three disabilities stratified by hospitalization status. Figure 3 shows the five symptoms for each outcome that demonstrated the strongest association. Dizziness (33% non-hospitalized, 39% hospitalized) was associated with all disability outcomes in both hospitalized and non-hospitalized groups. Heavy limbs (17%), dyspnea (49%), and tremors (8.5%) were associated with four of the five disability components in the non-hospitalized group, and heavy limbs (21%) with four of the five disability components in the hospitalized group (Tables S9.1-S9.10).

**Figure 3.**
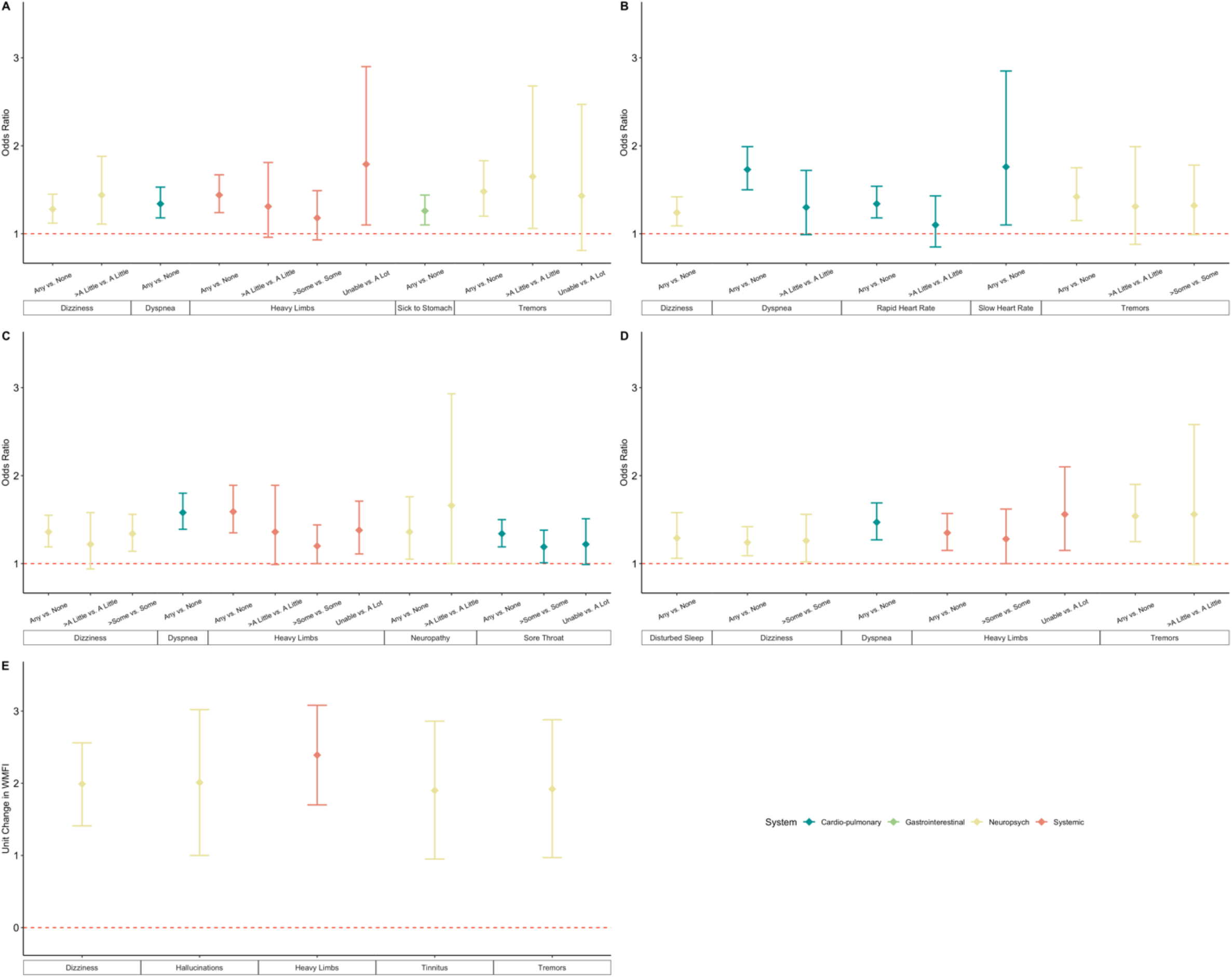
Association between symptoms and symptom severity at the time of initial COVID-19 illness and A: difficulty in walking a quarter of a mile; B: difficulty in walking up ten stairs; C: difficulty in doing light housework; D: difficulty in doing heavy housework; and E: level of mental fatigue. In each plot, the five symptoms with the strongest overall associations are shown. All models are adjusted for age, gender, body mass index, calendar period of infection, activity level prior to infection and vaccination status.

### Vaccination Status

We evaluated the effect of vaccination on long-COVID disability restricted to vaccine status *prior to* infection to reduce reverse causality from those unable to get a vaccine due to disability/health concerns. Individuals who received two mRNA doses/one adenoviral dose had a median time of approximately six months (IQR: 3.5, 8.5 months) between last dose and infection, while individuals who had 3+ mRNA/two adenoviral doses had two months (IQR: 1, 3 months) between last dose and infection. Vaccination was protective against development of mobility disability, IADL, and mental fatigue as compared to unvaccinated/one mRNA dose (Table 2: Model 1; Table S5.1-S5.2). We were not sufficiently powered to evaluate the interaction of time between vaccination and infection to account for potential waning vaccine effectiveness. In separate models, we adjusted for hospitalization (probably on the causal pathway between vaccination and disability); those who had 3+ mRNA/two adenoviral doses prior to their infection had a significantly decreased mobility disability, IADL, and mental fatigue compared to those who were unvaccinated/one mRNA dose (Table 2: Model 2; Table S5.1-S5.2). The additional IV analyses provided evidence of a protective effect of vaccination on mobility disability (Table 2: Model 3; Table S5.3-S5.5). While there was some protection afforded with respect to IADL disability, the estimate lacked precision, and there was no evidence of effect for mental fatigue.

**Table 2:**
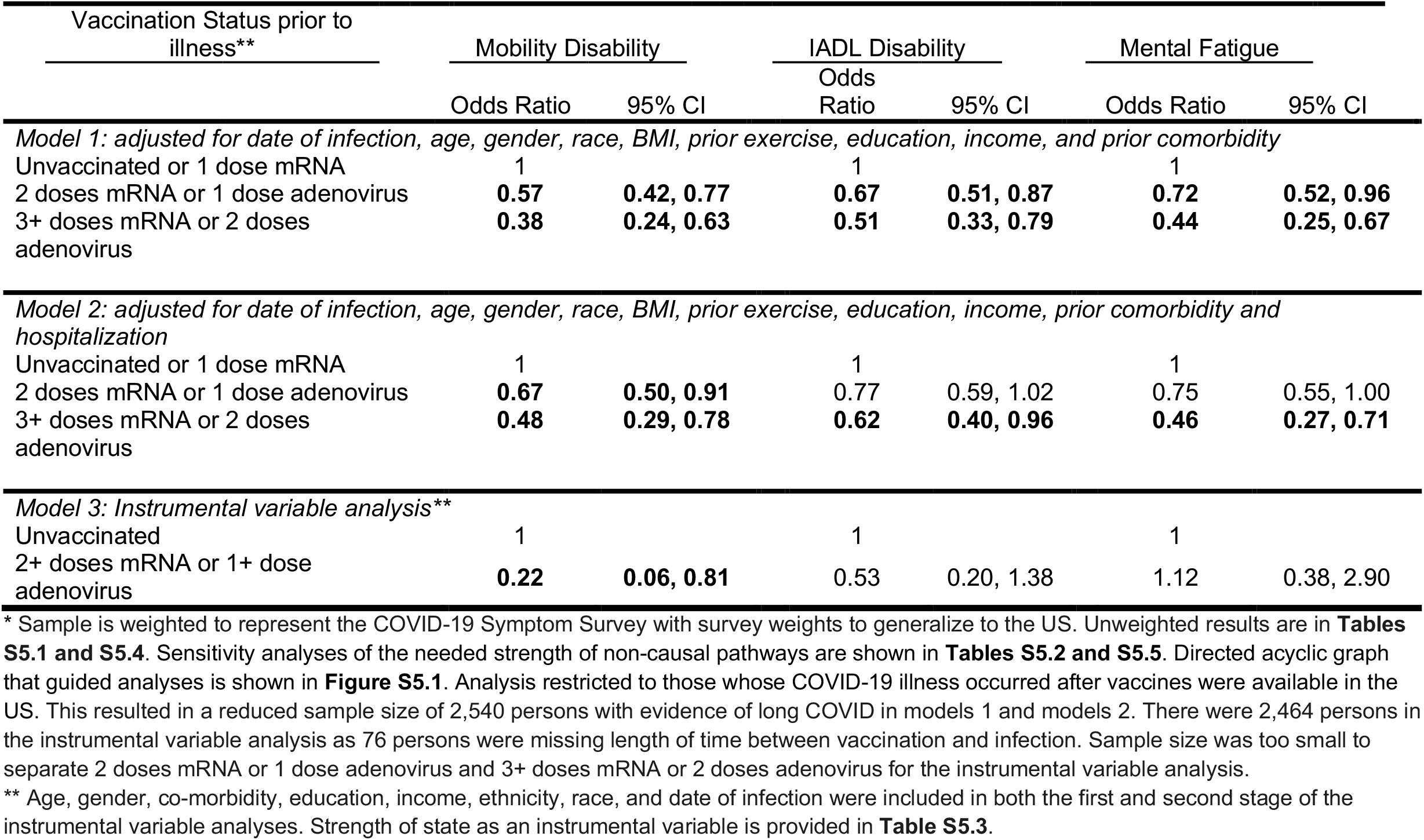
Effect of vaccination status prior to COVID-19 illness on disability*

## Discussion

In this large and heterogeneous sample from across the US, nearly two-thirds of persons with long-COVID classified as having some disability, a burden that was nearly 11 times that of those with resolved-COVID. Moreover, we identified that 1% were critically physically disabled or unable to function and 5% had critically severe mental fatigue. This has the potential to translate to millions of new disabilities in adults in the US. Considering that 30-90 million people are expected to have long-COVID (10-30% of cases), we might expect 300,000-900,000 people (1% of long-COVID) to be physically incapacitated, and 900,000-2.7 million (3% of long-COVID) to have critically severe mental fatigue. While it is reassuring that those with critically severe disabilities appear to be seeking necessary medical care (reflected by high levels of new physician diagnoses), these disabilities will likely compromise the ability of many to work or care for themselves and others, extending the long-term societal and economic impacts of COVID-19.

Our data was consistent with prior reports suggesting that hospitalization during acute illness is the strongest driver of COVID-19-associated disability. However, we also demonstrate that 60% of non-hospitalized individuals experience at least one long-COVID associated disability and 5.5% were classified with at least one critical disability. Although we observed some associations with age, gender, race, and pre-existing comorbidities with long-COVID disabilities, they contrasted with the risk factors of male gender, age, and comorbidities that have been consistently associated with COVID-19 severity and hospitalization. Essentially, these data suggest that factors predictive of hospitalization or severity of initial infection cannot be used to predict who will develop long-COVID-associated disability among those not requiring hospitalization.

While there was substantial heterogeneity in the presenting symptoms of acute COVID-19 in those with long-COVID, there were some patterns that emerged among those with incident disabilities. Among non-hospitalized individuals, dizziness, tremors, heavy limbs, and dyspnea were consistently associated with increased occurrence and sometimes severity of disability. Even among those hospitalized, dizziness and heavy limbs were related to all types of disability and severity. Dizziness and tremors are neurologic outcomes that may represent multiple disease paths including disruption of the vestibular system and dysautonomia, and postural orthostatic tachycardia syndrome (POTS). Associations of dizziness and tremors with imbalance may also explain observed associations with mobility/IADL disability.^20,21^ The sensation of heavy limbs is often attributable to venous insufficiency or peripheral arterial disease. This incident symptom may represent the effect of SARS-CoV-2 on the cardiovascular system with alterations to microvascular perfusion on the peripheral limbs, thromboembolism, and endothelial injury. While the high burden of physician diagnoses particularly among the most severely disabled group suggests that many are seeking clinical care, our data reinforces the need for those with early lingering symptoms to seek medical care that considers careful neurologic, vestibular, cardiovascular, and/or pulmonary evaluations. Additionally, there may be opportunities for the early administration of medications like corticosteroids^21^ or consideration of anti-platelet regimens and physical therapy, although it remains unclear whether early intervention will alleviate long-term impacts.

Additional clarity is needed on whether vaccination reduces disability risk associated with long-COVID. We observed over a 50% reduction in odds for all three disabilities among those with 3+ mRNA/two adenoviral doses and a more modest reduction in odds for those with two mRNA/one adenoviral dose. Some of this effect may be mediated through protection from severe COVID-19 resulting in hospitalization leading to disability. The strong reduction of mobility disability persisted in the two or more mRNA doses/one or more adenoviral dose IV analysis that is robust to measured or unmeasured confounding (Supplement Methods). With increasing viral positivity in the US, the window of complete vaccination (initial doses + booster) prior to infection is closing. However, these data support that vaccines may afford some protection from disabilities, as well as the intended purpose of attenuation of severe infection and hospitalization.

The cross-sectional design prevented us from exploring the impact of vaccination after infection on the resolution and/or exacerbation of COVID-19 associated disabilities. Comprehensive longitudinal studies are needed to explore the association of timing and dose of vaccination as well as number and timing of repeat infections on COVID-19 disability and changes over time. In our sample, we observed a high burden of all disabilities among those with long-COVID. While our sample may be skewed towards individuals reporting long-COVID, it was beneficial to identify common patterns in the long-COVID group, and especially those with disability. We included those with resolved-COVID and no-COVID for comparisons, acknowledging that the pandemic, outside of infection, was stressful and for some traumatic, and may contribute independently to disability. We also lack representation in demographic groups that were especially impacted by the pandemic, and thus we may underestimate the true burden of long-COVID disability across race/ethnicity, socio-economic, and community factors; however, this was minimized in the vaccine analysis by weighting the study sample to the COVID-19 Symptom Survey sample weights which increases the ability to generalize these results to the US population. Nevertheless, representativeness of these analyses depends on the differences in distribution of effect modifiers in our sample as compared to the general population (Supplement Methods and Figures S6.1-S6.3). The use of a study questionnaire provided us the opportunity to capture the heterogeneity in initial and continued symptoms but is subject to recall bias especially when the time of infection was far from enrollment in the study. However, this approach allowed us to be more inclusive of the spectrum of COVID-19 and long-COVID symptoms than studies that use electronic health records that rely on information from those seeking medical care only.

In conclusion, we observed a high burden of physical and mental disability among individuals who are experiencing long-COVID, which may have long term consequences for not only these individuals but society-at-large. Long-term follow-up will be required to understand how and for whom these disabilities will resolve or worsen over time. In the meantime, specialized care is needed for the growing population of persons with long-COVID to reduce the burden and impact of disability.

## Data Availability

All data produced in the present study are available upon reasonable request to the authors

## Author Contributions

Drs. Lau, Mehta and Duggal had full access to all of the data in the study and take responsibility for the integrity of the data and the accuracy of the data analysis.

Concept and Design: Lau, Mehta, Duggal

Statistical Analysis: Lau, Ni, Yenoykan

Acquisition, analysis or interpretation of data : All Authors

Drafting of the manuscript: Lau, Wenz, Mehta, Duggal

Critical revision of the manuscript for important intellectual content: All authors

Administrative, technical or material support: Wenz, Coggiano, Ni

## Conflict of Interest Disclosures

We have no conflicts to disclose.

## Funding/Support

This study was supported by the Johns Hopkins University COVID-19 Research Response Program and in part by Johns Hopkins University Center for AIDS Research (P30AI094189), which is supported by the following NIH Co-Funding and Participating Institutes and Centers: NIAID, NCI, NICHD, NHLBI, NIDA, NIA, NIGMS, NIDDK, NIMHD. The content is solely the responsibility of the authors and does not necessarily represent the official views of the NIH.

## Notes

### Competing Interest Statement

The authors have declared no competing interest.

### Author Declarations

The study was approved by the Johns Hopkins Bloomberg School of Public Health Institutional Review Board and all participants provide informed consent.

