## Supplementary Materials for "Physical and mental health disability associated with long-COVID: Baseline results from a US nationwide cohort"

### Supplementary Material


1) Study Design Schematic
2) Case Definitions for Woods Mental Fatigue Inventory
3) Inverse Odds Weight Estimation
4) Descriptive Analyses of Symptoms on Disability
5) Vaccination
    a. Standard Regression
    b. Instrumental Variable Analysis
        i. State as an Instrumental Variable
        ii. Results of both Unweighted and Weighted IV analyses
    c. Regression Based Analysis vs. IV Analysis
6) Representativeness
7) Venn Diagram of Disability Outcomes
8) Logistic regression analysis of non-symptom factors associated with each disability.
9) Tables of Sequential Logistic Model with each component of the disabilities
    a. Non-Hospitalized
        i. Walking a quarter of a mile
        ii. Climbing 10 stairs
        iii. Doing heavy housework
        iv. Doing light housework
        v. Woods Mental Fatigue Inventory
    b. Hospitalized
        i. Walking a quarter of a mile
        ii. Climbing 10 stairs
        iii. Doing heavy housework
        iv. Doing light housework
        v. Woods Mental Fatigue Inventory
10) Physician Diagnoses between Infection and Survey among those with Long-COVID
11) References

**1. Study Design Schematic:** In supplemental figure 1, we highlight the study flow of how we arrived at the sample sizes in each of the analyses.

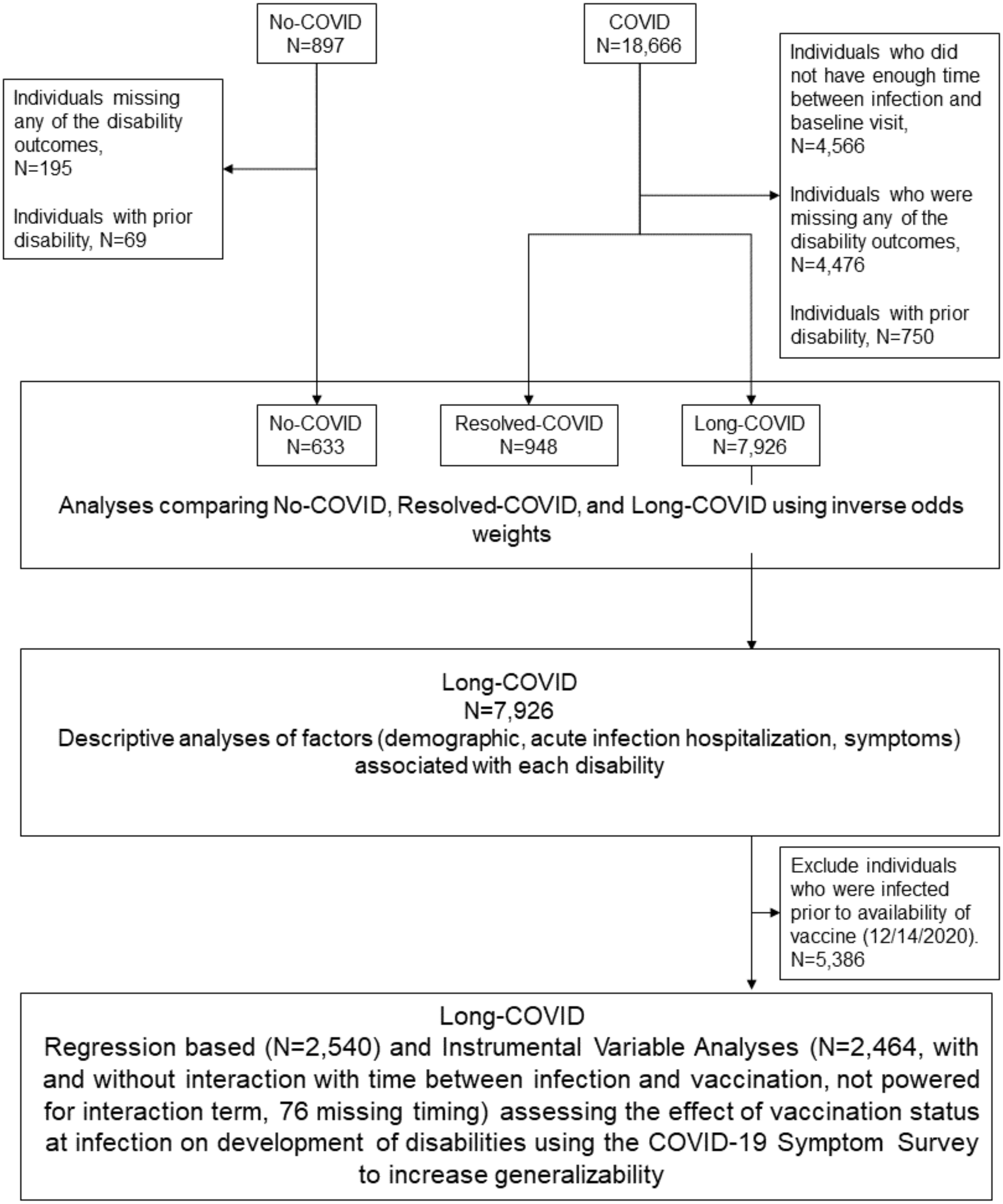


**Figure S1.1.** Study design Schematic

**2. Case Definition for Wood Mental Fatigue Inventory**

There are no established cutoffs for the Wood Mental Fatigue Inventory (WMFI). Previously, this validated instrument was used to define brain fog and more recently mental fatigue among individuals hospitalized for COVID-19 [1,2]. In this study, mental fatigue was defined as having a score > 20 on the WMFI (range 0-36 points). While a standard cutoff for mental fatigue is not defined, among healthy individuals, those in professions likely to experience greater degrees of stress (e.g., nurses and doctors) reported a mean score near 10, compared to a mean score of 5 for athletes and administrative staff [3]. Individuals with chronic fatigue syndrome reported a mean score of 19.[4] Considering these previously used values, we used a cutoff based on the 90th percentile of our non-COVID sample, which was a score of 20.

**3. Inverse Odds Weight Estimation**

Inverse odds weighting allows for the standardization of covariates. In this case, we wanted to ensure that those with no-COVID and resolved-COVID had distributions like long-COVID. We included the following variables in estimation of the inverse odds weights: diabetes, cardiovascular disease, history of heart attack, congestive heart failure, high blood pressure, high cholesterol, history of stroke, autoimmune disorders, hepatitis C, asthma, chronic lung disease, chronic kidney disease, cancer, depression, being pregnant, overweight, or obese, anxiety or other mental health condition, and chronic or acute Lyme disease.

To estimate the inverse odds weights, we used the machine learning approach, super learner, which is a stacked generalization ensemble method that uses multiple candidate methods for estimating the probability of being in the long-COVID group.[5–8] This algorithm performs at least as well as the best single candidate method included[5] and protects from overfitting through the use a k-fold cross-validation. The candidate methods included in the super learner were a mix of regression based approaches and non-parametric tree based methods: 1) elastic net with hyper parameters to include ridge (alpha=0), lasso (alpha=1), and a mix between these two (alpha=0.5), 2) multivariate adaptive splines with hyperparameters for the degree of interaction of 1, 2, or 3, 3) random forest with the number of candidate variables to try at each split of 2, 3, and 7 variables, 4) support vector machines with C-constant regularization term of 1, 5, or 10 with either a radial basis kernel or Laplacian kernel, and 5) gradient boosted machines with either 500 or 1000 trees, a max depth of 2 or 4, and shrinkage of 0.01 or 0.1. This gives a combination of 23 candidate algorithms across each combination of parameters.

**4. Symptom Analyses**

We assessed the association of the presenting symptoms during the acute phase of infection with disability severity. Rather than focus on the domain of mobility and IADL, we examined the association with the components of the domain (mobility disability: difficulty in walking ¼ mile, difficulty in climbing 10 stairs; IADL: difficulty in heavy housework, difficulty in light housework). We used a relaxed LASSO approach with both multinomial and logistic models. The reason for relaxed LASSO is that LASSO by itself is not the best variable selection approach in all situations[9]; however, a relaxed LASSO is good at identifying variables in both low and high signal to noise ratio settings.[10] More specifically, the LASSO estimate is weighted with the least squares estimate creating a tension in which the estimates are being shrunk by LASSO with the weights relaxing the shrinkage. Once variable sets were identified, we evaluated both a multinomial and sequential logistic model on the variable sets, and the model with the lowest Akaike's Information Criterion (AIC) was selected. We used a sequential logistic model given the nested structure of the variable for severity and because it is not subject to the assumption of "independence of irrelevant alternatives" like the multinomial.[11] We chose not to use the ordinal logistic model because it assumes proportional odds and those assumptions were expected to be violated.

**5. Vaccination**

Making causal inferences in the estimation of the effect of vaccination on disability requires exchangeability between those who were and were not vaccinated. We used a standard regression approach as well as an instrumental variable analysis to estimate whether vaccination had impacts on mobility disability, IADL disability, and mental fatigue. This overall approach was chosen due to the different assumptions in each of the analyses. For these analyses we restricted to individuals who were infected on or after December 14, 2020, when vaccine became available to some individuals.

As COVID-Long data comes from a survey where recruitment was done via social-media and word of mouth, the distribution of US states across the sample is not reflective of the general population. For the IV analyses we present a weighted estimate based upon the likelihood of the observation being in 'COVID-Long' versus 'The COVID-19 Symptom Survey'. The COVID-19 Symptom Survey was an opportunity to collect data among Facebook users and was done in partnership with Carnegie Mellon University. On average there were 30,533 persons reporting COVID-19 in the past 14 days per wave with 13 waves of data. To make the COVID-19 Symptom Survey generalizable to the US, a sampling design and bias correction was developed to estimate survey weights.[12] Therefore, to control for potential selection bias in the IV analyses we estimated inverse probability of selection weights.

To create the weights from the COVID-19 Symptom Survey, we used "wave 8" data which corresponds to February and March 2021 (to reflect overlap with the COVID-Long survey). We limited the COVID-19 Symptom Survey to individuals who developed COVID-19. Variables of interest from the COVID-19 Symptom Survey were harmonized with the COVID-Long data. After harmonizing these variables, "wave 8" data was imputed given the amount of missing data in both the COVID-19 Symptom Survey and COVID-Long. To produce inverse probability of selection weights, we regressed being in 'COVID-Long' versus 'The COVID-19 Symptom Survey' based upon characteristics that may be related to development of long-COVID and disability. These included the following variables: gender, age, education, race/ethnicity, state, prior comorbidity (cancer - other than skin, heart disease, high blood pressure, asthma, chronic lung disease, kidney disease, diabetes, autoimmune problems, obesity), symptoms (fever, cough, sore throat, shortness of breath, stuffy or runny nose, chills, tiredness or exhaustion, chest pressure, nausea or vomiting, diarrhea, muscle or joint aches, headaches, loss of smell and taste, eye problems, new or developing symptoms (cough, tiredness or exhaustion, chest pressure, nausea or vomiting, diarrhea, headache, chills, loss of smell and taste, muscle or joint aches, eye problems, being pregnant, and recent anxiety). The data was imputed 50 times using "mice" package in R. We estimated the probability of being in the sample using the weighted logistic model with the above set of the covariates, where the survey weights from the COVID-19 Symptom Survey were used to allow for the generalization of results to the general population in the US [ref]. For the individuals in COVID-Long, we used a weight of 1. The average weights were obtained by averaging across 50 sets of the stabilized weights. Because models with inverse probability weights are subject to instability, it is also well established that the variance is quite high.[13] We therefore utilized a smooth-IPW approach with these weights following a recent method.[13] ***Therefore, the inverse probability weights should account for both 1) selection into 'COVID-Long' as compared to 'The COVID-19 Symptom Survey'; and 2) generalizability to the US.***

**5a. Standard Regression**

For conclusions to have a causal interpretation (i.e., causal inference) several assumptions must be met. These assumptions include: 1) exchangeability (i.e., all confounders are controlled for such that there are no unmeasured confounders or residual confounding that remains)[14]; any unmeasured confounders or residual confounding that remains is of no importance given the level of precision; 2) consistency of treatment[15]; specifically, we have assumed that even if there is variation in vaccination between mRNA and adenoviral vector vaccine, they would result in similar treatment effects such that we do not have to assume treatment variation by type of vaccine; and 3) positivity (i.e., that individuals have some positive probability of either being vaccinated or unvaccinated).[16] The following DAG helped to guide this analysis. Note, we have implicitly controlled for having COVID-19 and having long-COVID in our analysis as vaccination may impact if individuals may be infected. We colored the edge from vaccination to disability which is the effect that we are trying to estimate with a different color for ease of reading.

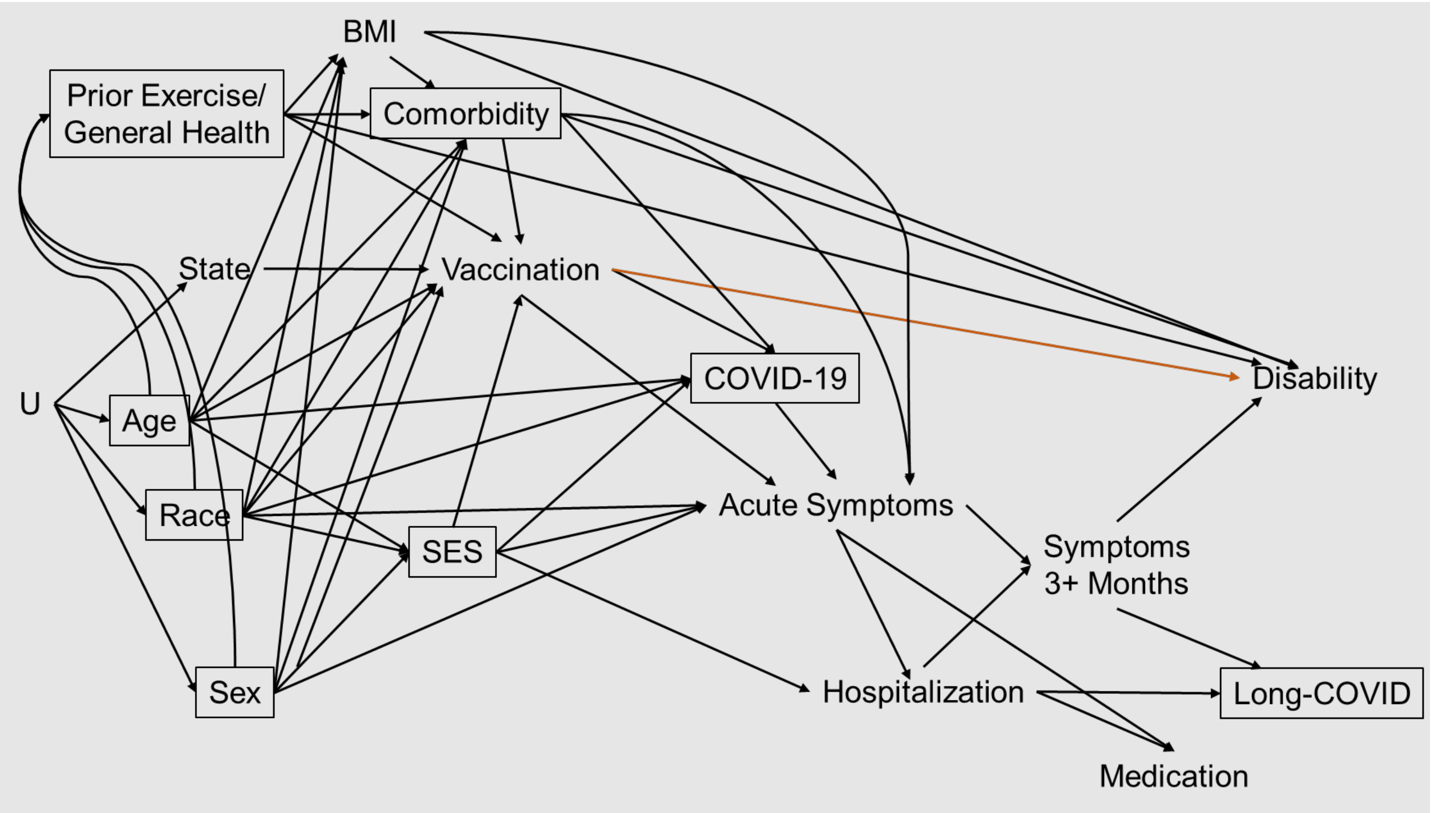


**Figure S5.1.** Directed acyclic graph of the relationship of vaccination to disability due to COVID-19

Unweighted regression analyses results are as follows (weighted analysis included in main text Table 2):

**Table S5.1:** Unweighted regression of the effect of vaccination on disability without (Model 1) and with (Model 2) hospitalization

| Vaccination Status prior to illness** | Mobility Disability | | IADL Disability | | Mental Fatigue | |
|---|---|---|---|---|---|---|
| | Odds Ratio | 95% CI | Odds Ratio | 95% CI | Odds Ratio | 95% CI |
| *Model 1: adjusted for date of infection, age, gender, race, BMI, prior exercise, education, income, and prior comorbidity* | | | | | | |
| Unvaccinated or 1 dose mRNA | 1 | | 1 | | 1 | |
| 2 doses mRNA or 1 dose adenovirus | **0.62** | **0.47, 0.83** | **0.75** | **0.58, 0.98** | 0.77 | 0.59, 1.02 |
| 3+ doses mRNA or 2 doses adenovirus | **0.44** | **0.27, 0.71** | **0.59** | **0.38, 0.92** | **0.48** | **0.29, 0.78** |
| *Model 2: adjusted for date of infection, age, gender, race, BMI, prior exercise, education, income, prior comorbidity and hospitalization* | | | | | | |
| Unvaccinated or 1 dose mRNA | 1 | | 1 | | 1 | |
| 2 doses mRNA or 1 dose adenovirus | **0.72** | **0.54, 0.97** | 0.87 | 0.66, 1.14 | 0.81 | 0.61, 1.07 |
| 3+ doses mRNA or 2 doses adenovirus | **0.53** | **0.32, 0.88** | 0.72 | 0.46, 1.12 | **0.51** | **0.31, 0.83** |

As unmeasured confounding remains a potential issue, the following table describes the E-values for the relationships described in Table 2 for models 1 and 2. E-value is a measure of the necessary strength of an unmeasured confounder's association with exposure and association with outcome that is required to change inference.[17,18] These results suggest that the totality of unmeasured confounding for mobility disability for 3+ doses mRNA or 2 doses adenovirus (model 1) needs to have a measure of association of 2.09 with the exposure and 2.09 with mobility disability. In other words, this suggests that a fairly strong confounder would be required to nullify the estimate and an unmeasured confounder would require an association of 1.46 with exposure and with mobility disability to have the OR confidence interval include 1.

**Table S5.2:** Sensitivity of weighted results in Table 2 (main text) to unmeasured confounding. The E-value is the minimum strength of the unmeasured confounder with vaccination and minimum strength of unmeasured confounder with outcome of disability to move the results to the null (OR=1). The E-Value CI is the same as the E-value but the strength needed to move the confidence interval to include 1.

| Vaccination Status prior to illness** | Mobility Disability | | IADL Disability | | Mental Fatigue | |
|---|---|---|---|---|---|---|
| | E-value | E-Value CI | E-value | E-Value CI | E-value | E-Value CI |
| *Model 1: adjusted for date of infection, age, gender, race, BMI, and prior comorbidity* | | | | | | |
| Unvaccinated or 1 dose mRNA | 1 | | 1 | | 1 | |
| 2 doses mRNA or 1 dose adenovirus | 1.98 | 1.54 | 1.74 | 1.35 | 1.64 | 1.17 |
| 3+ doses mRNA or 2 doses adenovirus | 2.63 | 1.83 | 2.15 | 1.50 | 2.38 | 1.74 |
| *Model 2: adjusted for date of infection, age, gender, race, BMI, and prior comorbidity and hospitalization* | | | | | | |
| Unvaccinated or 1 dose mRNA | 1 | | 1 | | 1 | |
| 2 doses mRNA or 1 dose adenovirus | 1.74 | 1.27 | 1.53 | 1 | 1.58 | 1 |
| 3+ doses mRNA or 2 doses adenovirus | 2.24 | 1.52 | 1.86 | 1.17 | 2.31 | 1.66 |

**5b. Instrumental Variable Analysis**

An instrumental variable (IV) approach was also used to estimate the effect of vaccination on each disability as different assumptions are made to interpret a causal effect of vaccination. The advantage of an instrumental variable analysis is that meeting the exchangeability assumption does not depend on accounting for all confounders. Therefore, the analysis is protected against misspecification of the model by not including confounders due to omission whether this is due to the confounder not being measured or the relationship with the confounder having not yet been elucidated. However, other assumptions must be met to have a valid causal inference. The variable(s) that is being used as the instrument must fulfill three criteria: 1) the instrument must affect the exposure or be a descendent of a cause of the exposure, 2) the instrument affects the outcome only through the exposure, and 3) the instrument does not share a common cause with the outcome[19,20,21].

The IV analysis approach is the two-stage residual inclusion (2SRI) model for binary outcomes.[22] By including the residual from the first stage model, the residual acts as a control function that helps to minimize potential bias due to the nonlinear outcome model. [23] In doing so, the 2SRI model estimates the conditional causal odds ratio. In general, IV analyses estimate the local average treatment effect which therefore results as the interpretation of the effect of vaccination among the compliers.[20] Similarly, this conditional causal odds [24] ratio is the effect among the compliers. For estimating the 2SRI model, we used ivglm in the R package ivtools. However, because we include weights in the IV analysis, we bootstrapped the analysis 1000 times to obtain the 95% confidence intervals.

**State as an Instrumental Variable**

From the DAG (Figure S5.1), we identified US state as a potential instrument. Outside of the accounted for individual factors, we believe that the association between state and disability is mediated through vaccination. Additionally, state is associated with vaccination. However, there is little guidance on the strength of an instrumental variable for binary exposure. In a recent paper on IV analyses, Hughes et al. generated a simulation that relied on the $R^2$ statistic with a strong instrument being approximately 0.39 and a moderate instrument having a $R^2$ of 0.045 (supplement in [25]). Therefore, we provide McFadden's pseudo-$R^2$. We chose this particular $R^2$ as it specifically attempts to measure the amount of explained variation as the –2*log(likelihood) is equivalent to the sum of squared errors when the model is an ordinary least squares. Thus, McFadden's pseudo-$R^2$, which is based upon the log(likelihood) for a model with additional covariates as compared to the reduced model, provides the proportion reduction in the –2* log(likelihood) paralleling that of an ordinary least squares model.[26] We estimated the strength of the instrument in a simple intercept and instrument (state) model as well as in the presence of other covariates. The other covariates included age, gender, prior comorbidity, education, income, race, and date of infection.

Table S5.3 details the pseudo-R and partial pseudo-R with and without the inverse probability weights estimated with 'The COVID-19 Symptom Survey' data. The results suggest that state is a moderate strength instrument for the instrumental variable analysis and therefore provides protection against bias that can be introduced with weak instruments.[27]

**Table S5.3:** Strength of the variable state as an instrumental variable with and without inverse probability weights using the COVID-19 Symptom Survey

| **Model** | **With Inverse Probability of Selection Weights** | **McFadden's Pseudo-$R^2$** |
|---|---|---|
| Intercept + **State** vs. Intercept only | No | 0.043 |
| Intercept + **State** vs. Intercept only model | Yes | 0.011 |
| Intercept + Additional Covariates + **State** vs. Intercept + Additional Covariates | No | 0.054 |
| Intercept + Additional Covariates + **State** vs. Intercept + Additional Covariates | Yes | 0.073 |

*Additional Covariates included: age, gender, prior comorbidity, education, income, ethnicity, race, and date of infection

**Results of both Unweighted and Weighted IV analyses**

**Table S5.4:** Weighted and unweighted results on the instrumental variable analyses estimating the effect of vaccination (2+ mRNA doses or 1+ adenoviral vector dose as compared to unvaccinated or those with 1 mRNA dose) *

| | Mobility Disability | | IADL | | Mental Fatigue | |
|---|---|---|---|---|---|---|
| | OR | 95% CI | OR | 95% CI | OR | 95% CI |
| No Weights | **0.311** | **(0.153, 0.822)** | 0.483 | (0.236, 1.132) | 1.260 | (0.483, 2.790) |
| Weights included in second stage | **0.263** | **(0.126, 0.805)** | 0.442 | (0.212, 1.188) | 1.122 | (0.415, 2.933) |
| Weights included in both first and second stage | **0.221** | **(0.059, 0.812)** | 0.532 | (0.201, 1.380) | 1.121 | (0.380, 2.904) |

*Second stage model included: age, gender, prior comorbidity, education, income, ethnicity, race, and date of infection plus the predicted vaccination and the residual from the first stage model

The overall inference remains the same regardless of the inclusion of weights as shown in Table S5.4. Similar to the regression-based approach, we can measure the strength of a non-causal pathway between vaccination and the outcome of mobility disability. Given that this is an IV analysis, this would not be an unmeasured confounder but the relationship due to selection bias or information bias in which the number of paths equate to having some factor relationship with exposure and this factor with outcome (i.e., selection and exposure; and selection and outcome). The relationship between selection and exposure as well as selection and outcome each may be made of several causal pathways but overall sum to the E-value. The E-value and E-value CI are shown in the following table and suggest a fairly strong relationship between exposure and selection and selection and outcome to change inference.

**Table S5.5:** Summary of non-causal pathways that would be required to move the estimate to an OR of 1.0 or the value required to have the 95% confidence interval include 1.0 (E-value CI)

| | **Mobility Disability** | |
|---|---|---|
| | E-value | E-value CI |
| No Weights | 2.99 | 1.44 |
| Weights included in second stage | 3.31 | 1.47 |
| Weights included in both first and second stage | 3.68 | 1.46 |

**5.c Regression Based Analysis vs. IV Analysis**
While both analyses result in similar qualitative inferences (i.e., protective effect of vaccination on mobility disability), the measure of effect being estimated is not actually the same. The regression-based estimates are estimating the conditional causal odds ratio in the entire sample whereas the IV analysis is estimating the conditional causal odds ratio among the compliers. Therefore, it is not surprising that the conditional odds ratio of the IV analysis is stronger in magnitude as compared to the regression-based approach. Furthermore, the definition of "vaccination" is different between the two analyses. In the regression-based approach the sample size is adequate to estimate the number of doses of vaccination as 2 mRNA or 1 adenovirus and 3+ mRNA or 2 doses of adenoviral vector vaccine as compared to unvaccinated or 1 mRNA vaccination. However, with increased uncertainty in an IV analysis, we collapsed the two vaccination categories as we are underpowered to split into multiple categories.

**6. Representativeness**
The goal of a study sample being representative is to help make inference in the target population.
A study is representative if the distribution of the effect modifiers is the same as the target population. Therefore, it is not whether the demographics of a group of individuals (e.g., those of low, middle, or high social economic status) is included in the sample with the same distribution. Rather, if the demographics or other factor are an effect modifier of the relationships being assessed and if the distributions are equivalent. For example, if a study asked whether essential personnel are at higher risk of being infected than non-essential personnel and the distribution of sex (male/female) is not the same as the general population the results may still be generalized to the general population There has been no evidence for a difference in *susceptibility* by sex for SARS-CoV-2 infection as multiple studies have shown similar incidences of infection[28]. Therefore, there is no reason to expect sex to modify the relationship of essential worker status on the incidence of SARS-CoV-2. Thus, the average effect of essential personnel status on incident infection among females would generalize to males and vice versa. The distribution of sex in the study sample could be entirely female or entirely male and still represent the relationship of essential personnel status on incident infection in the target population.

In this study, the sample recruitment was done primarily through social media. It is likely that our sample skews towards individuals having more severe disease including likelihood of having long-COVID and disability. However, having a higher potential for selection into the sample based on the outcomes does not necessarily invalidate comparisons. For instance, comparing individuals who are vaccinated as compared to those are not vaccinated on the incidence of disability *may have* the same DAG as earlier (Figure S.6.1.) with the addition of selection into the sample.

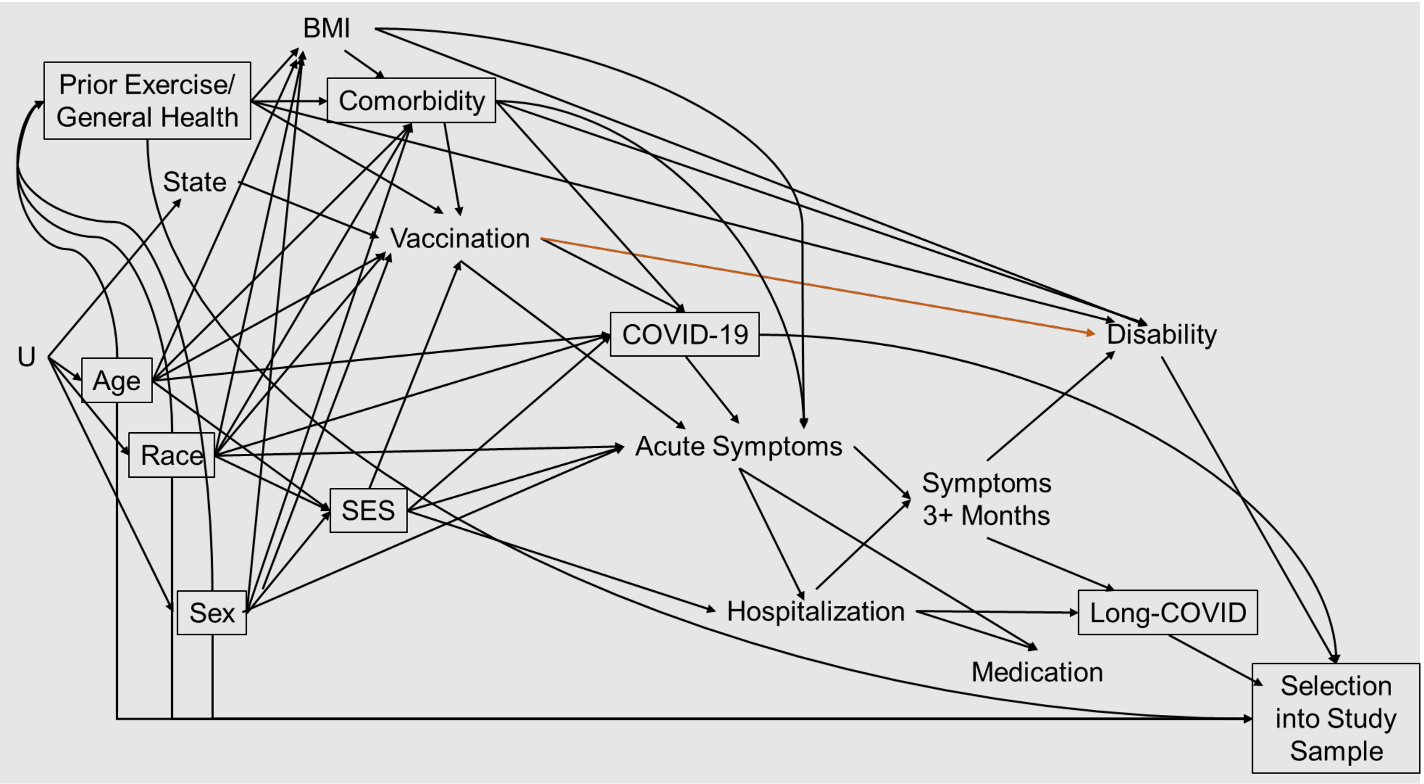


**Figure S6.1** Directed acyclic graph representing individuals being selected into sample

Given that age, sex, race, social economic status (via education and income), co-morbidity, are adjusted for and that the sample is restricted to those with COVID-19 and those that have long-COVID, we can reduce the DAG to the following schematic (Figure S.6.2), which is analogous to one for a case-control study (Figure S.6.3). Therefore, estimation of the causal odds ratio remains unbiased in samples in which the selection into the study is based upon the outcome status.[29] Whether the sample is representative in terms of effect modifiers is likely unknown given that the knowledge of the relationship of disability among those with long-COVID is currently limited. Furthermore, even if the distribution of the effect modifiers is not the same, we may be able to generalize the qualitative inferences (e.g., increased or decreased risk) and not the overall magnitude of the treatment effect depending on proportion of groups that have an opposing qualitative effect from the treatment effect and the over-representations (i.e., proportion of sample) of these groups in the sample.

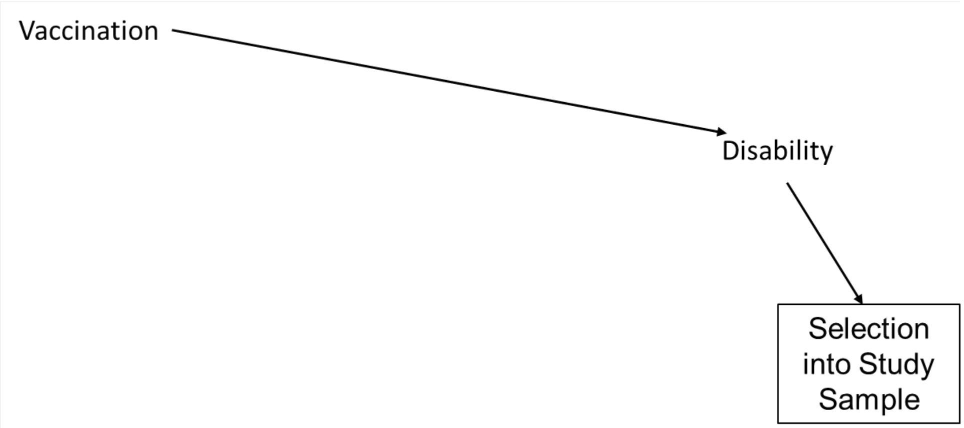


**Figure S6.2** Resulting directed acyclic graph between vaccination and selection in study removing blocked pathways

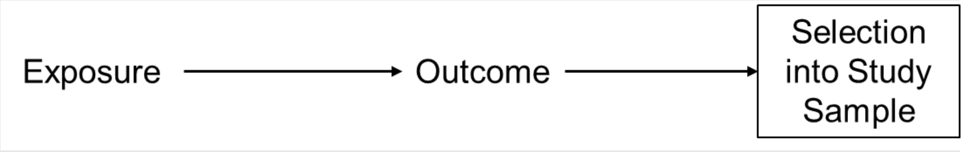


**Figure S6.3** Directed acyclic graph for case-control study

Furthermore, we attempted to minimize differences in potentially relevant distributions of characteristics in our sample and that of the ‘The COVID-19 Symptom Survey’ using inverse probability selection weights. Additionally, the survey weights provided by the ‘The COVID-19 Symptom Survey’ allows for generalization to the broader US population. The magnitude of the effects estimated as well as the weights suggest that the inferences of the estimates may be generalized and if not the estimates, then the qualitative inference of protective effect of vaccination should remain robust and generalized to the US population. For the association of symptoms during acute infection, we controlled for multiple factors that could be potential confounders of the relationship according to our DAG. However, we would caution interpretation of the relationship between symptom and disability as being causal as the symptoms themselves may represent several different disease paths. For example, dizziness and tremors may reflect underlying disease paths such as hypertension, dehydration, postural orthostatic tachycardia syndrome (POTS), vestibular neuritis, vertigo, multiple sclerosis, and neurodegenerative disease. Thus, this information should be used as informative for both identifying individuals that may be at high risk for disability as well as focusing further research.

#### 7. Venn Diagram of Disability Outcomes

A.

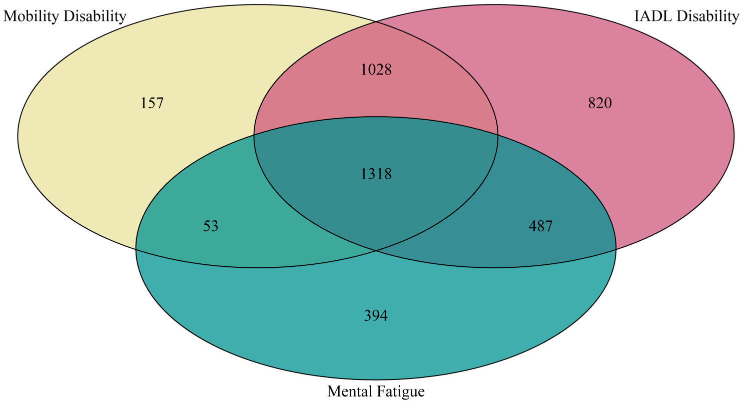


B.

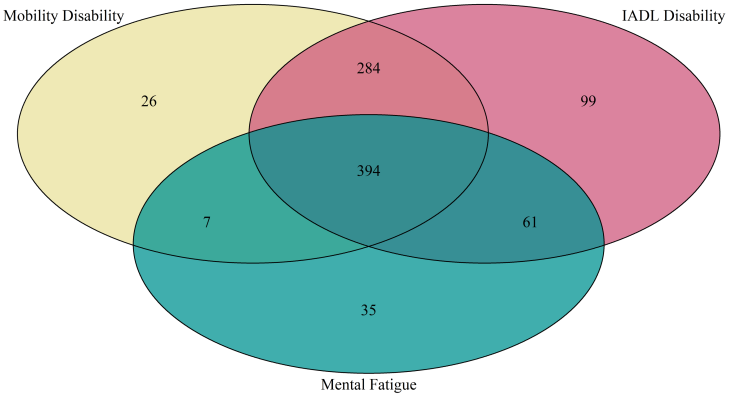


**Figure S7**. Venn diagram of disability outcomes (mobility disability, IADL disability and mental fatigue) stratified by hospitalization status during acute infection. Panel A includes 4,115 persons who were not hospitalized at the time of their initial illness and had at least one disability. Panel B includes 906 persons who were hospitalized at the time of their initial illness and had at least one disability. Thirteen persons with missing data on hospitalization status were excluded.

**8. Logistic regression models of non-symptom factors associated with each disability.**

**Table S8.1.** Associations between non-symptom factors and mobility disability defined as either at least some difficulty walking a quarter of a mile or climbing ten stairs among persons with long-COVID*

| | All participants (n=7926) | | Non-Hospitalized participants (n=6870) | | Hospitalized participants (n=1049) | |
|---|---|---|---|---|---|---|
| | Unadjusted OR (95% CI) | Adjusted OR (95% CI) | Unadjusted OR (95% CI) | Adjusted OR (95% CI) | Unadjusted OR (95% CI) | Adjusted OR (95% CI) |
| Age (per 10 years) | 1.25 (1.20, 1.29) | 1.26 (1.21, 1.31) | 1.24 (1.19, 1.29) | 1.28 (1.23, 1.33) | 1.06 (0.95, 1.17) | 1.15 (1.03, 1.29) |
| Gender | | | | | | |
| Male | 1 | 1 | 1 | 1 | 1 | 1 |
| Female | 1.53 (1.34, 1.75) | 1.80 (1.56, 2.08) | 1.66 (1.43, 1.94) | 1.76 (1.50, 2.07) | 1.88 (1.37, 2.57) | 1.88 (1.35, 2.63) |
| Transgender, Genderqueer, or Non-Binary | 1.51 (1.03, 1.05) | 1.59 (1.06, 2.39) | 1.49 (1.00, 2.24) | 1.60 (1.05, 2.44) | 2.31 (0.51, 10.49) | 1.77 (0.38, 8.33) |
| Race | | | | | | |
| White | 1 | 1 | 1 | 1 | 1 | 1 |
| Non-white | 1.19 (1.00, 1.43) | 1.22 (1.00, 1.48) | 1.07 (0.87, 1.31) | 1.20 (0.97, 1.49) | 1.29 (0.81, 2.06) | 1.31 (0.80, 2.14) |
| Multi-race | 1.20 (0.93, 1.56) | 1.29 (0.98, 1.69) | 1.26 (0.95, 1.66) | 1.34 (1.00, 1.79) | 0.92 (0.44, 1.79) | 0.91 (0.42, 2.01) |
| Education | | | | | | |
| Any Post-Graduate Studies | 1 | 1 | 1 | 1 | 1 | 1 |
| Less than or Equal to High School | 2.28 (1.87, 2.79) | 1.91 (1.54, 2.37) | 2.16 (1.73, 2.70) | 1.96 (1.55, 2.48) | 1.96 (1.15, 3.34) | 1.48 (0.83, 2.64) |
| Some College, Associates Degree, or Technical Degree | 2.11 (1.88, 2.36) | 1.68 (1.48, 1.90) | 1.97 (1.74, 2.23) | 1.66 (1.45, 1.89) | 2.12 (1.53, 2.94) | 1.69 (1.19, 2.41) |
| Bachelor's Degree | 1.06 (0.94, 1.18) | 1.06 (0.94, 1.19) | 1.06 (0.94, 1.20) | 1.06 (0.94, 1.20) | 1.02 (0.73, 1.43) | 0.99 (0.62, 1.25) |
| Income (per year) | | | | | | |
| < $15,000 | 1.57 (1.26, 1.95) | 1.65 (1.30, 2.10) | 1.43 (1.12, 1.81) | 1.53 (1.18, 1.98) | 4.00 (1.75, 9.14) | 3.68 (1.55, 8.73) |
| ≥ $15,000 to < $25,000 | 2.16 (1.73, 2.69) | 1.96 (1.55, 2.48) | 2.07 (1.63, 2.63) | 1.98 (1.53, 2.55) | 2.20 (1.20, 4.06) | 1.86 (0.98, 3.52) |
| ≥ $25,000 to < $35,000 | 2.07 (1.70, 2.52) | 1.83 (1.48, 2.26) | 1.96 (1.58, 2.43) | 1.75 (1.40, 2.19) | 3.46 (1.76, 6.80) | 2.96 (1.46, 5.99) |
| ≥ $35,000 to < $50,000 | 1.71 (1.46, 2.00) | 1.49 (1.26, 1.76) | 1.64 (1.39, 1.95) | 1.47 (1.23, 1.76) | 2.04 (1.29, 3.21) | 1.68 (1.05, 2.70) |
| ≥ $50,000 to < $75,000 | 1.51 (1.33, 1.70) | 1.39 (1.22, 1.58) | 1.42 (1.25, 1.63) | 1.33 (1.16, 1.53) | 2.21 (1.53, 3.21) | 1.83 (1.24, 2.70 |
| ≥ $75,000 | 1 | 1 | 1 | 1 | 1 | 1 |
| Body mass index (per kg/$m^2$) | 1.04 (1.04, 1.05) | 1.03 (1.02, 1.04) | 1.04 (1.03, 1.05) | 1.03 (1.02, 1.04) | 1.03 (1.01, 1.05) | 1.02 (1.00, 1.04) |
| Prior comorbidity | 1.60 (1.44, 1.78) | 1.27 (1.21, 1.31) | 1.62 (1.45, 1.82) | 1.32 (1.17, 1.50) | 1.14 (0.83, 1.56) | 0.88 (0.62, 1.25) |
| Prior activity level | | | | | | |
| Insufficient activity | 1 | 1 | 1 | 1 | 1 | 1 |
| Moderate activity | 0.94 (0.75, 1.18) | 1.05 (0.82, 1.34) | 0.95 (0.74, 1.22) | 1.05 (0.81, 1.37) | 1.00 (0.55, 1.81) | 1.13 (0.61, 2.12) |
| Active | 1.09 (0.92, 1.29) | 1.49 (1.24, 1.79) | 1.12 (0.93, 1.36) | 1.47 (1.21, 1.80) | 1.15 (0.74, 1.79) | 1.51 (0.94, 2.43) |
| Hospitalization | 3.55 (3.09, 4.08) | 3.07 (2.65, 3.55) | NA | NA | NA | NA |

* 7 persons refused to answer whether they were hospitalized or not during their acute infection

**Table S8.2.** Associations between non-symptom factors and IADL disability defined as having at least some difficulty with heavy housework or light housework among persons with long-COVID*

| | All participants (n=7926) | | Non-Hospitalized participants (n=6870) | | Hospitalized participants (n=1049) | |
|---|---|---|---|---|---|---|
| | Unadjusted OR (95% CI) | Adjusted OR (95% CI) | Unadjusted OR (95% CI) | Adjusted OR (95% CI) | Unadjusted OR (95% CI) | Adjusted OR (95% CI) |
| Age (per 10 years) | 1.31 (1.27, 1.36) | 1.34 (1.29, 1.39) | 1.32 (1.27, 1.38) | 1.37 (1.31, 1.43) | 0.98 (0.87, 1.10) | 1.07 (0.93, 1.22) |
| Gender | | | | | | |
| Male | 1 | 1 | 1 | 1 | 1 | 1 |
| Female | 1.58 (1.39, 1.79) | 1.88 (1.64, 2.15) | 1.66 (1.45, 1.91) | 1.81 (1.56, 2.09) | 2.22 (1.57, 3.13) | 2.23 (1.54, 3.21) |
| Transgender, Genderqueer, or Non-Binary | 1.78 (1.17, 2.72) | 2.00 (1.29, 3.09) | 1.79 (1.16, 2.77) | 2.04 (1.31, 3.20) | 2.53 (0.33, 19.6) | 1.77 (0.22, 14.2) |
| Race | | | | | | |
| White | 1 | 1 | 1 | 1 | 1 | 1 |
| Non-white | 1.02 (0.86, 1.23) | 1.09 (0.90, 1.32) | 0.95 (0.78, 1.16) | 1.10 (0.89, 1.35) | 0.94 (0.57, 1.57) | 1.07 (0.62, 1.83) |
| Multi-race | 1.19 (0.91, 1.54) | 1.30 (0.99, 1.72) | 1.15 (0.87, 1.52) | 1.27 (0.95, 1.70) | 1.78 (0.62, 5.13) | 1.97 (0.65, 5.96) |
| Education | | | | | | |
| Any Post-Graduate Studies | 1 | 1 | 1 | 1 | 1 | 1 |
| Less than or Equal to High School | 1.80 (1.46, 2.22) | 1.64 (1.31, 2.05) | 1.66 (1.32, 2.08) | 1.62 (1.28, 2.06) | 1.85 (1.00, 3.40) | 1.73 (0.90, 3.35) |
| Some College, Associates Degree, or Technical Degree | 1.90 (1.69, 2.13) | 1.64 (1.45, 1.86) | 1.73 (1.53, 1.95) | 1.58 (1.38, 1.80) | 2.50 (1.70, 3.68) | 2.26 (1.48, 3.43) |
| Bachelor's Degree | 1.09 (0.98, 1.22) | 1.13 (1.00, 1.26) | 1.07 (0.96, 1.20) | 1.11 (0.98, 1.25) | 1.29 (0.88, 1.89) | 1.26 (0.85, 1.88) |
| Income (per year) | | | | | | |
| < $15,000 | 1.18 (0.95, 1.47) | 1.32 (1.03, 1.67) | 1.09 (0.87, 1.38) | 1.28 (0.99, 1.64) | 2.86 (1.10, 7.41) | 2.11 (0.78, 5.68) |
| ≥ $15,000 to < $25,000 | 1.86 (1.47, 2.34) | 1.78 (1.39, 2.28) | 1.71 (1.34, 2.19) | 1.75 (1.35, 2.28) | 2.69 (1.19, 6.08) | 2.05 (0.88, 4.77) |
| ≥ $25,000 to < $35,000 | 1.63 (1.33, 2.00) | 1.54 (1.24, 1.92) | 1.57 (1.27, 1.95) | 1.53 (1.22, 1.92) | 2.13 (1.02, 4.44) | 1.67 (0.77, 3.62) |
| ≥ $35,000 to < $50,000 | 1.39 (1.19, 1.63) | 1.28 (1.08, 1.51) | 1.35 (1.14, 1.59) | 1.30 (1.09, 1.55) | 1.56 (0.93, 2.62) | 1.25 (0.73, 2.16) |
| ≥ $50,000 to < $75,000 | 1.36 (1.20, 1.53) | 1.32 (0.95, 1.82) | 1.29 (1.13, 1.47) | 1.29 (1.12, 1.47) | 2.14 (1.37, 3.36) | 1.69 (1.06, 2.70) |
| ≥ $75,000 | 1 | 1 | 1 | 1 | 1 | 1 |
| Body mass index (per kg/m$^2$) | 1.02 (1.02, 1.03) | 1.01 (1.00, 1.01) | 1.02 (1.01, 1.02) | 1.01 (1.00, 1.01) | 1.02 (1.00, 1.04) | 1.01 (0.99, 1.03) |
| Prior comorbidity | 1.56 (1.41, 1.72) | 1.37 (1.23, 1.53) | 1.53 (1.37, 1.70) | 1.38 (1.23, 1.56) | 1.39 (0.98, 1.98) | 1.29 (0.87, 1.93) |
| Prior activity level | | | | | | |
| Insufficient activity | 1 | 1 | 1 | 1 | 1 | 1 |
| Moderate activity | 1.23 (0.99, 1.53) | 1.35 (1.07, 1.71) | 1.22 (0.96, 1.55) | 1.32 (1.03, 1.69) | 1.55 (0.81, 2.97) | 1.80 (0.91, 3.56) |
| Active | 1.55 (1.32, 1.83) | 2.05 (1.72, 2.46) | 1.15 (1.33, 1.91) | 1.27 (1.64, 2.41) | 1.91 (1.19, 3.05) | 1.97 (0.65, 5.96) |
| Hospitalization | 3.50 (2.99, 4.10) | 3.14 (2.67, 3.70) | NA | NA | NA | NA |

* 7 persons refused to answer whether they were hospitalized or not during their acute infection

**Table S8.3.** Associations between non-symptom factors and mental fatigue defined as greater or equal to a score of 20 on the Woods Mental Fatigue Inventory*

| | All participants (n=7926) | | Non-Hospitalized participants (n=6870) | | Hospitalized participants (n=1049) | |
|---|---|---|---|---|---|---|
| | Unadjusted OR (95% CI) | Adjusted OR (95% CI) | Unadjusted OR (95% CI) | Adjusted OR (95% CI) | Unadjusted OR (95% CI) | Adjusted OR (95% CI) |
| Age (per 10 years) | 0.96 (0.93, 1.00) | 0.96 (0.92, 1.00) | 0.97 (0.93, 1.01) | 0.99 (0.95, 1.03) | 0.79 (0.72m 0.87) | 0.83 (0.75, 0.92) |
| Gender | | | | | | |
| Male | 1 | 1 | 1 | 1 | 1 | 1 |
| Female | 1.38 (1.20, 1.58) | 1.41 (1.22, 1.63) | 0.69 (0.59, 0.80) | 1.43 (1.22, 1.68) | 1.38 (1.01, 1.89) | 1.29 (0.93, 1.79) |
| Transgender, Genderqueer, or Non-Binary | 1.74 (1.19, 2.16) | 1.56 (1.05, 2.30) | 1.70 (1.13, 2.55) | 1.56 (1.03, 2.36) | 2.37 (0.72, 7.76 | 1.63 (0.47, 5.59) |
| Race | | | | | | |
| White | 1 | 1 | 1 | 1 | 1 | 1 |
| Non-white | 1.03 (0.86, 1.25) | 1.00 (0.83, 1.22) | 1.03 (0.83, 1.27) | 1.05 (0.84, 1.30) | 0.84 (0.55, 1.27) | 0.88 (0.57, 1.36) |
| Multi-race | 1.29 (0.99, 1.68) | 1.23 (0.94, 1.60) | 1.36 (1.02, 1.80) | 1.29 (0.97, 1.72) | 0.96 (0.48, 1.95) | 0.79 (0.38, 1.67) |
| Education | | | | | | |
| Any Post-Graduate Studies | 1 | 1 | 1 | 1 | 1 | 1 |
| Less than or Equal to High School | 1.63 (1.33, 2.00) | 1.52 (1.23, 1.89) | 1.61 (1.28, 2.03) | 1.57 (1.24, 1.99) | 1.38 (0.86, 2.22) | 1.30 (0.78, 2.19) |
| Some College, Associates Degree, or Technical Degree | 1.88 (1.67, 2.11) | 1.71 (1.51, 1.94) | 1.83 (1.61, 2.08) | 1.72 (1.50, 1.96) | 1.78 (1.31, 2.40) | 1.62 (1.17, 2.26) |
| Bachelor's Degree | 1.13 (1.00, 1.27) | 1.10 (0.98, 1.24) | 1.10 (0.97, 1.25) | 1.09 (0.96, 1.24) | 1.30 (0.93, 1.81) | 1.14 (0.80, 1.61) |
| Income (per year) | | | | | | |
| < $15,000 | 1.60 (1.28, 2.00) | 1.29 (1.02, 1.63) | 1.58 (1.24, 2.01) | 1.31 (1.02, 1.68) | 1.73 (0.94, 3.19) | 1.25 (0.65, 2.42) |
| ≥ $15,000 to < $25,000 | 1.80 (1.44, 2.24) | 1.48 (1.18, 1.87) | 1.64 (1.28, 2.09) | 1.41 (1.09, 1.81) | 2.36 (1.36, 4.11) | 1.92 (1.07, 3.44) |
| ≥ $25,000 to < $35,000 | 1.46 (1.19, 1.79) | 1.18 (0.96, 1.46) | 1.49 (1.20, 1.86) | 1.23 (0.98, 1.55) | 1.25 (0.74, 2.11) | 1.00 (0.57, 1.76) |
| ≥ $35,000 to < $50,000 | 1.75 (1.49, 2.04) | 1.49 (1.27, 1.75) | 1.71 (1.44, 2.03) | 1.48 (1.24, 1.76) | 1.79 (1.19, 2.71) | 1.66 (1.07, 2.56) |
| ≥ $50,000 to < $75,000 | 1.37 (1.21, 1.55) | 1.24 (1.09, 1.41) | 1.38 (1.20, 1.58) | 1.28 (1.11, 1.47) | 1.23 (0.88, 1.71) | 1.01 (0.71, 1.44) |
| ≥ $75,000 | 1 | 1 | 1 | 1 | 1 | 1 |
| Body mass index (per kg/m$^2$) | 1.01 (1.00, 1.02) | 1.00 (0.99, 1.01) | 1.01 (1.01, 1.02) | 1.00 (0.99, 1.01) | 1.00 (0.99, 1.02) | 1.00 (0.98, 1.02) |
| Prior comorbidity | 1.40 (1.26, 1.56) | 1.35 (1.21, 1.52) | 1.44 (1.28, 1.62) | 1.39 (1.23, 1.58) | 0.98 (0.73, 1.32) | 1.08 (0.78, 1.51) |
| Prior activity level | | | | | | |
| Insufficient activity | 1 | 1 | 1 | 1 | 1 | 1 |
| Moderate activity | 0.96 (0.75, 1.22) | 1.01 (0.79, 1.29) | 1.00 (0.77, 1.30) | 1.04 (0.80, 1.36) | 0.79 (0.43, 1.44) | 0.86 (0.46, 1.60) |
| Active | 1.27 (1.06, 1.52) | 1.42 (1.18, 1.71) | 1.21 (0.99, 1.47) | 1.34 (1.10, 1.65) | 1.83 (1.18, 2.85) | 1.84 (1.16, 2.92) |
| Hospitalization | 1.85 (1.62, 2.11) | 1.78 (1.55, 2.04) | NA | NA | NA | NA |

* 7 persons refused to answer whether they were hospitalized or not during their acute infection

### 9. Tables of Sequential Logistic Model with each Component of each Disability

**Table S9.1**. Associations between symptoms at initial COVID-19 illness and self-reported severity of difficulty in walking a quarter of a mile among participants who were not hospitalized at the time of initial illness*

| Symptom** | Any vs. None OR (95% CI) | >A Little vs. A Little OR (95% CI) | >Some vs. Some OR (95% CI) | Unable vs. A Lot OR (95% CI) | Prevalence of Symptom or Factor | Number of Times Selected by Algorithms**** |
|---|---|---|---|---|---|---|
| Shortness of Breath | **1.47 (1.27, 1.69)** | | | | 0.49 | **1.00** |
| Heavy Limbs | **1.35 (1.15, 1.57)** | | **1.28 (1.00, 1.62)** | **1.56 (1.15, 2.10)** | 0.17 | **1.00** |
| Dizziness | **1.24 (1.09, 1.42)** | | **1.26 (1.02, 1.56)** | | 0.33 | **1.00** |
| Tremors | **1.54 (1.25, 1.90)** | 1.56 (0.99, 2.58) | | | 0.08 | 0.67 |
| Disturbed Sleep | **1.29 (1.06, 1.58)** | | | | 0.12 | 0.67 |
| Pneumonia | 1.25 (0.98, 1.58) | | | | 0.06 | 1.00 |
| Tightness in Chest | **1.23 (1.07, 1.41)** | | | | 0.44 | 0.67 |
| Neuropathy | 1.22 (0.97, 1.54) | 1.65 (0.95, 3.02) | | | 0.08 | 0.67 |
| Sick to Stomach | **1.21 (1.05, 1.39**) | | | | 0.23 | 0.67 |
| Insomnia | 1.21 (0.98, 1.84) | | | | 0.12 | 0.67 |
| Slow Heart Rate | 1.19 (0.74, 1.90) | | | | 0.01 | 0.67 |
| Sore Throat | **1.18 (1.05, 1.34)** | | | | 0.48 | 0.67 |
| Fast Heart Rate | **1.16 (1.01, 1.33)** | | | | 0.33 | 0.67 |
| Eye Problems | 1.11 (0.88, 1.39) | 1.48 (0.84, 2.75) | | | 0.08 | 0.67 |
| Muscle Weakness | 1.10 (0.95, 1.28) | | | | 0.49 | 0.67 |
| Cough | 1.07 (0.95, 1.22) | | | | 0.47 | 0.67 |
| Alopecia | 1.07 (0.90, 1.28) | | | | 0.12 | 0.67 |
| Fever | 0.92 (0.78, 1.08) | | | | 0.14 | 0.67 |
| Lack of Energy | | **1.63 (1.09, 2.38)** | | | 0.86 | 0.67 |
| Fever >100.4 | | | **1.24 (1.01, 1.52)** | | 0.39 | 0.67 |
| Runny Nose | 0.92 (0.81, 1.05) | | | | 0.38 | 0.67 |
| Loss of Appetite | 0.91 (0.80, 1.03) | | | | 0.45 | 0.67 |
| Loss of Smell | **0.84 (0.74, 0.95)** | | | | 0.49 | 0.67 |
| Altered Smell | **0.59 (0.46, 0.75)** | | | | 0.08 | 0.67 |

* Models were adjusted for age, gender, BMI, calendar period of infection, activity level prior to infection, prior comorbidity, and vaccination status
**Symptoms were selected utilizing a relaxed least absolute shrinkage and selection operator (LASSO) *** Only 7 persons were boosted and hence the wide confidence intervals **** Three methods for variable selection were used: 1) Random Forest, 2) Multinomial Model with relaxed LASSO, 3) Sequential logistic model with relaxed LASSO; For models with LASSO, the lambda (shrinkage) parameter and gamma (relaxing) parameter were chosen through 20-fold cross validation; Variables included in the model was determined by applying each variable set determined by algorithms and choosing model with the best fit by Akaike's Information Criterion

**Table S9.2.** Associations between symptoms at initial COVID-19 illness and self-reported severity of difficulty in walking up 10 stairs among participants who were not hospitalized at the time of initial illness*

| Symptom** | Any vs. None OR (95% CI) | >A Little vs. A Little**** OR (95% CI) | >Some vs. Some OR (95% CI) | Unable vs. A Lot OR (95% CI) | Prevalence of Symptom or Factor | Number of Times Selected by Algorithms*** |
|---|---|---|---|---|---|---|
| Shortness of Breath | **1.73 (1.50, 1.99)** | 1.30 (0.99, 1.72) | | | 0.49 | **1.00** |
| Dizziness | **1.24 (1.09, 1.42)** | | | | 0.33 | **1.00** |
| Slow Heart Rate | **1.76 (1.10, 2.85)** | | | | 0.01 | 0.67 |
| Tremors | **1.42 (1.15, 1.75)** | 1.31 (0.88, 1.99) | 1.32 (0.99, 1.78) | | 0.08 | 0.67 |
| Rapid Heart Rate | **1.34 (1.18, 1.54)** | 1.10 (0.85, 1.43) | | | 0.33 | 0.67 |
| Neuropathy | **1.32 (1.04, 1.66)** | | | | 0.08 | 0.67 |
| Heavy Limbs | **1.22 (1.05, 1.43)** | | **1.30 (1.02, 1.65)** | 1.55 (0.90, 2.65) | 0.17 | 0.67 |
| Muscle Weakness | **1.20 (1.02, 1.40)** | | | | 0.49 | 0.67 |
| Sick to Stomach | **1.19 (1.03, 1.37)** | 1.09 (0.82, 1.44) | | | 0.23 | 0.67 |
| Disturbed Sleep | 1.19 (0.98, 1.45) | 1.07 (0.73, 1.59) | | | 0.12 | 0.67 |
| Insomnia | 1.18 (0.96, 1.44) | 1.16 (0.79, 1.73) | | | 0.12 | 0.67 |
| Sore Throat | **1.15 (1.02, 1.29)** | 1.11 (0.88, 1.39) | | | 0.48 | 0.67 |
| Eye Problems | 1.14 (0.91, 1.44) | 1.22 (0.80, 1.90) | | | 0.08 | 0.67 |
| Cough | 1.13 (1.00, 1.28) | | | | 0.47 | 0.67 |
| Tightness in Chest | 1.10 (0.96, 1.26) | 1.02 (0.77, 1.34) | | | 0.44 | 0.67 |
| Pneumonia | 1.10 (0.87, 1.40) | 1.41 (0.89, 2.33) | | | 0.06 | 0.67 |
| Joint Aches | 1.07 (0.92, 1.23) | 1.20 (0.93, 1.54) | | | 0.43 | 0.67 |
| Tinnitus | 1.06 (0.85, 1.33) | | | | 0.08 | 0.67 |
| Muscle Aches | 1.05 (0.91, 1.23) | | | | 0.60 | 0.67 |
| Alopecia | 1.03 (0.86, 1.22) | 1.16 (0.82, 1.66) | | | 0.12 | 0.67 |
| Loss of Taste | | 1.26 (0.99, 1.60) | | | 0.44 | 0.67 |
| Joint Aches | | 1.20 (0.93, 1.54) | | | 0.43 | 0.67 |
| Skin Problems | | | | 2.28 (1.00, 4.84) | 0.05 | 0.67 |
| Fever | 0.89 (0.75, 1.04) | | | | 0.14 | 0.67 |
| Loss of Appetite | 0.88 (0.78, 1.00) | | | | 0.45 | 0.67 |
| Loss of Smell | **0.85 (0.76, 0.96)** | | | | 0.49 | 0.67 |
| Diarrhea | **0.82 (0.72, 0.94)** | 1.16 (0.89, 1.51) | | | 0.32 | 0.67 |
| Altered Smell | **0.62 (0.49, 0.78)** | **0.55 (0.37, 0.82)** | | | 0.08 | 0.67 |
| Chills | | 1.06 (0.83, 1.35) | | | 0.49 | 0.33 |
| Altered Taste | | **0.55 (0.37, 0.82)** | | | 0.09 | 0.33 |

* Models were adjusted for age, gender, BMI, calendar period of infection, activity level prior to infection, and vaccination status **Symptoms were selected utilizing a relaxed least absolute shrinkage and selection operator (LASSO)*** Three methods for variable selection were used: 1) Random

Forest, 2) Multinomial Model with relaxed LASSO, 3) Sequential logistic model with relaxed LASSO; For models with LASSO, the lambda (shrinkage) parameter and gamma (relaxing) parameter were chosen through 20-fold cross validation; Variables included in the model was determined by applying each variable set determined by algorithms and choosing model with the best fit by Akaike's Information Criterion**** Seizure was also selected for this model, but all individuals who had seizure also had more difficulty in climbing ten stairs so estimate is unstable.

**Table S9.3**. Associations between symptoms at initial COVID-19 illness and self-reported severity of difficulty in doing heavy housework among participants who were not hospitalized at the time of initial illness*

| Symptom** | Any vs. None OR (95% CI) | >A Little vs. A Little OR (95% CI) | >Some vs. Some OR (95% CI) | Unable vs. A Lot OR (95% CI) | Prevalence of Symptom or Factor | Number of Times Selected by Algorithms*** |
|---|---|---|---|---|---|---|
| Heavy Limbs | **1.59 (1.35, 1.89)** | 1.36 (0.99, 1.89) | 1.20 (1.00, 1.44) | **1.38 (1.11, 1.71)** | **0.17** | **1.00** |
| Shortness of Breath | **1.58 (1.39, 1.80)** | | | | 0.49 | **1.00** |
| Dizziness | **1.36 (1.19, 1.55)** | 1.22 (0.94, 1.58) | **1.34 (1.14, 1.56)** | | 0.33 | **1.00** |
| Sore Throat | **1.34 (1.19, 1.50)** | | **1.19 (1.01, 1.38)** | 1.22 (0.99, 1.51) | 0.48 | **1.00** |
| Eye Problems | 1.27 (0.99, 1.63) | 1.33 (0.83, 2.25) | **1.40 (1.09, 1.80)** | | 0.08 | **1.00** |
| Tremors | 1.27 (0.99, 1.64) | | **1.42 (1.11, 1.85)** | 1.24 (0.94, 1.63) | 0.08 | **1.00** |
| Neuropathy | **1.36 (1.05, 1.76)** | 1.66 (1.00, 2.93) | | | 0.08 | **1.00** |
| Tinnitus | **1.30 (1.02, 1.65)** | | | | 0.08 | **1.00** |
| Rapid Heart Rate | **1.28 (1.12, 1.47)** | 1.26 (0.97, 1.64) | | | 0.33 | **1.00** |
| Sick to Stomach | **1.27 (1.11, 1.47)** | 1.30 (0.99, 1.74) | | | 0.23 | **1.00** |
| Joint Aches | **1.23 (1.08, 1.39)** | | | | 0.43 | **1.00** |
| Difficulty Sleeping | **1.17 (1.02, 1.35)** | | | | 0.27 | **1.00** |
| Tightness in Chest | 1.14 (1.00, 1.29) | 1.24 (0.98, 1.59) | | | 0.44 | **1.00** |
| Pneumonia | | | | 1.23 (0.86, 1.73) | 0.06 | **1.00** |
| Loss of Appetite | 0.86 (0.76, 0.97) | | | | 0.45 | **1.00** |
| Slow Heart Rate | 1.47 (0.83, 2.71) | | | | 0.01 | 0.67 |
| Insomnia | 1.19 (0.97, 1.47) | | | | 0.12 | 0.67 |
| Disturbed Sleep | 1.19 (0.97, 1.46) | | | | 0.12 | 0.67 |
| Hallucinations | 1.19 (0.92, 1.55) | | | | 0.06 | 0.67 |
| Lack of Energy | 1.10 (0.93, 1.29) | 1.15 (0.83, 1.59) | | | 0.86 | 0.67 |
| Muscle Weakness | 1.08 (0.94, 1.24) | | | | 0.49 | 0.67 |
| Alopecia | 1.07 (0.89, 1.30) | 1.25 (0.86, 1.88) | | | 0.12 | 0.67 |
| Cough | 1.05 (0.93, 1.17) | | **1.18 (1.01, 1.37)** | 1.18 (0.96, 1.45) | 0.47 | 0.67 |
| Runny Nose | **0.89 (0.80, 1.00)** | | | **0.74 (0.60, 0.91)** | 0.38 | 0.67 |
| Loss of Smell | **0.77 (0.69, 0.86)** | | | **0.79 (0.64, 0.96)** | 0.49 | 0.67 |
| Altered Smell | **0.65 (0.52, 0.80)** | | | | 0.08 | 0.67 |
| Seizure | 0.36 (0.14, 1.01) | | | | 0.003 | 0.67 |

* Models were adjusted for age, gender, BMI, calendar period of infection, activity level prior to infection, and vaccination status. **Symptoms were selected utilizing a relaxed least absolute shrinkage and selection operator (LASSO). *** Three methods for variable selection were used: 1) Random Forest, 2) Multinomial Model with relaxed LASSO, 3) Sequential logistic model with relaxed LASSO; For models with LASSO, the lambda (shrinkage) parameter and gamma (relaxing) parameter were chosen through 20-fold cross validation; Variables included in the model was determined by applying each variable set determined by algorithms and choosing model with the best fit by Akaike's Information Criterion

**Table S9.4.** Associations between symptoms at initial COVID-19 illness and self-reported severity of difficulty in doing light housework among participants who were not hospitalized at the time of initial illness*

| Symptom** | Any vs. None OR (95% CI) | >A Little vs. A Little OR (95% CI) | >Some vs. Some OR (95% CI) | Unable vs. A Lot OR (95% CI) | Prevalence of Symptom or Factor | Number of Times Selected by Algorithm*** |
|---|---|---|---|---|---|---|
| Heavy Limbs | **1.44 (1.24, 1.67)** | 1.31 (0.96, 1.81) | 1.18 (0.93, 1.49) | **1.79 (1.10, 2.90)** | **0.17** | **1.00** |
| Tremors | **1.48 (1.20, 1.83)** | **1.65 (1.06, 2.68)** | | 1.43 (0.81, 2.47) | 0.08 | **1.00** |
| Shortness of Breath | **1.34 (1.18, 1.53)** | | | | 0.49 | **1.00** |
| Dizziness | **1.28 (1.12, 1.45)** | **1.44 (1.11, 1.88)** | | | 0.33 | **1.00** |
| Sick to Stomach | **1.26 (1.10, 1.44)** | | | | 0.23 | **1.00** |
| Slow Heart Rate | **1.65 (1.03, 2.68)** | | | | 0.01 | 0.67 |
| Rapid Heart Rate | **1.29 (1.13, 1.47)** | | | | 0.33 | 0.67 |
| Neuropathy | **1.28 (1.01, 1.62)** | | 1.18 (0.86, 1.63) | | 0.08 | 0.67 |
| Sore Throat | **1.27 (1.13, 1.42)** | | | | 0.48 | 0.67 |
| Tightness in Chest | **1.20 (1.06, 1.38)** | | | | 0.44 | 0.67 |
| Tinnitus | 1.20 (0.96, 1.50) | | | | 0.08 | 0.67 |
| Disturbed Sleep | 1.16 (0.95, 1.41) | | | | 0.12 | 0.67 |
| Insomnia | 1.15 (0.94, 1.40) | | 1.32 (0.99, 1.76) | | 0.12 | 067 |
| Eye Problems | 1.13 (0.89, 1.42) | | | 1.71 (0.95, 3.01) | 0.08 | 0.67 |
| Hypersomnia | 1.12 (0.92, 1.35) | | | | 0.12 | 0.67 |
| Joint Aches | 1.10 (0.97, 1.24) | | | | 0.43 | 0.67 |
| Difficulty Sleeping | 1.09 (0.95, 1.26) | | | | 0.27 | 0.67 |
| Hallucinations | 1.05 (0.84, 1.32) | | | | 0.06 | 0.67 |
| Alopecia | | **1.54 (1.05, 2.33)** | 1.21 (0.93, 1.57) | | 0.12 | 0.67 |
| Pain in Teeth | | | 1.25 (0.95, 1.64) | | 0.12 | 0.67 |
| Runny Nose | | | 0.77 (0.63, 0.93) | | 0.38 | 0.67 |
| Loss of Smell | 0.81 (0.72, 0.90) | | | | 0.49 | 0.67 |
| Altered Smell | 0.63 (0.50, 0.79) | | | | 0.08 | 0.67 |
| Muscle Weakness | | | 1.12 (0.90, 1.39) | | 0.49 | 0.33 |

*Models were adjusted for age, gender, BMI, calendar period of infection, activity level prior to infection, and vaccination status. **Symptoms were selected utilizing a relaxed least absolute shrinkage and selection operator (LASSO). *** Three methods for variable selection were used: 1) Random Forest, 2) Multinomial Model with relaxed LASSO, 3) Sequential logistic model with relaxed LASSO; For models with LASSO, the lambda (shrinkage) parameter and gamma (relaxing) parameter were chosen through 20-fold cross validation; Variables included in the model was determined by applying each variable set determined by algorithms and choosing model with the best fit by Akaike's Information Criterion

**Table S9.5.** Associations between symptoms at initial COVID-19 illness and self-reported severity of mental fatigue as measured by the Woods Mental Fatigue Inventory among participants who were not hospitalized at the time of initial illness*

| Symptom** | Unit Change in WMFI | 95% CI | Prevalence of Symptom or Factor | Proportion of Times Selected By Algorithms*** |
|---|---|---|---|---|
| Heavy Limbs | **2.39** | **1.70, 3.08** | 0.17 | **1.00** |
| Hallucinations | **2.01** | **1.00, 3.02** | 0.06 | **1.00** |
| Dizziness | **1.99** | **1.41, 2.56** | 0.33 | **1.00** |
| Tremors | **1.92** | **0.97, 2.88** | 0.08 | **1.00** |
| Tinnitus | **1.90** | **0.95, 2.86** | 0.08 | **1.00** |
| Hypersomnia | **1.00** | **0.20, 1.79** | 0.12 | **1.00** |
| Joint Aches | **1.38** | **0.83, 1.92** | 0.44 | **1.00** |
| Eye Problems | 0.92 | -0.08, 1.91 | 0.08 | **1.00** |
| Muscle Weakness | **0.88** | **0.26, 1.51** | 0.49 | **1.00** |
| Shortness of Breath | **1.45** | **0.92, 1.99** | 0.49 | 0.50 |
| Disturbed Sleep | **1.28** | **0.43, 2.14** | 0.12 | 0.50 |
| Slow Heart Rate | 1.12 | -0.93, 3.18 | 0.01 | 0.50 |
| Sore Throat | **1.06** | **0.58, 1.54** | 0.48 | 0.50 |
| Insomnia | **1.05** | **0.20, 1.90** | 0.12 | 050 |
| Lack of Energy | **1.01** | **0.29, 1.73** | 0.86 | 0.50 |
| Loss of Taste | **1.00** | **0.24, 1.75** | 0.45 | 0.50 |
| Headache | **0.77** | **0.22, 1.32** | 0.71 | 0.50 |
| Difficulty Sleeping | **0.75** | **0.13, 1.37** | 0.27 | 0.50 |
| Sick to Stomach | 0.49 | -0.10, 1.08 | 0.23 | 0.50 |
| Alopecia | 0.44 | -0.33, 1.21 | 0.12 | 0.50 |
| Fever | 0.34 | -0.31, 1.00 | 0.14 | 0.50 |
| Rapid Heart Rate | 0.29 | -0.29, 0.86 | 0.33 | 0.50 |
| Cough | -0.30 | -0.80, 0.20 | 0.48 | 0.50 |
| Chills | -0.35 | -0.86, 0.17 | 0.49 | 0.50 |
| Loss of Smell | **-1.02** | **-1.78, -0.26** | 0.49 | 0.50 |
| Altered Smell | **-1.05** | **-1.96, -0.15** | 0.08 | 0.50 |
| Skin Problems | **-1.48** | **-2.62, -0.34** | 0.05 | 0.50 |
| Vaccination | | | | |
| Fully | -0.99 | (-2.00, 0.02) | 0.08 | |
| Boosted | **-1.94** | **(-3.67, -0.22)** | 0.02 | |

* Models were adjusted for age, gender, BMI, calendar period of infection, activity level prior to infection, and vaccination status

**Symptoms were selected utilizing a relaxed least absolute shrinkage and selection operator (LASSO)

*** Two methods for variable selection were used: 1) Random Forest, 2) Gaussian generalized linear model with relaxed LASSO; For model with LASSO, the lambda (shrinkage) parameter and gamma (relaxing) parameter were chosen through 20-fold cross validation; Variables included in the model was determined by applying each variable set determined by algorithms and choosing model with the best fit by Akaike's Information Criterion

**Table S9.6.** Associations between symptoms at initial COVID-19 illness and self-reported severity of difficulty in walking a quarter of a mile among participants who were hospitalized at the time of initial illness*

| | **Any vs. None** | **>A Little vs. A Little** | **>Some vs. Some** | **Unable vs. A Lot** | **Prevalence of Symptom or Factor** | **Proportion of Times Selected by Algorithms**** |
|---|---|---|---|---|---|---|
| **Symptom**** | **OR (95% CI)** | **OR (95% CI)** | **OR (95% CI)** | **OR (95% CI)** | | |
| Heavy Limbs | **1.93 (1.34, 2.80)** | | | | 0.21 | **1.00** |
| Dizziness | **1.41 (1.04, 1.92)** | 1.65 (0.80, 3.48) | | | 0.39 | **1.00** |
| Shortness of Breath | 1.14 (0.83, 1.57) | 1.66 (0.86, 3.17) | | | 0.72 | 0.67 |
| Rapid Heart Rate | | 2.05 (0.98, 4.44) | | | 0.45 | 0.67 |
| Tightness in Chest | | 1.16 (0.53, 2.52) | | | 0.58 | 0.67 |
| Joint Aches | 1.21 (0.91, 1.61) | | | | 0.50 | 0.33 |
| Cough | 1.18 (0.90, 1.56) | | | | 0.58 | 0.33 |
| Chills | | 1.64 (0.53, 3.17) | | | 058 | 0.33 |
| Sick to Stomach | | **0.48 (0.25, 0.93)** | | | 0.33 | 0.33 |
| Skin Problems | | 0.48 (0.18, 1.36) | | | 0.08 | 0.33 |
| Altered Smell | | **0.37 (0.15, 0.96)** | | | 0.09 | 0.33 |
| Fever >100.4 | **0.64 (0.49, 0.84)** | | | | 0.53 | 0.33 |

* Models were adjusted for age, gender, BMI, calendar period of infection, activity level prior to infection, prior comorbidity, and vaccination status
** Three methods for variable selection were used: 1) Random Forest, 2) Multinomial Model with relaxed LASSO, 3) Sequential logistic model with relaxed LASSO; For models with LASSO, the lambda (shrinkage) parameter and gamma (relaxing) parameter were chosen through 20-fold cross validation; Variables included in the model was determined by applying each variable set determined by algorithms and choosing model with the best fit by Akaike's Information Criterion. Final model was sequential logit model with variables selected from sequential logit model with relaxed LASSO.
*** Only four individuals were boosted with no variability in outcome and therefore a stable estimate could not be obtained
**** Only two individuals were boosted with no variability in the outcome and therefore a stable estimate could not be obtained

**Table S9.7**. Associations between symptoms at initial COVID-19 illness and self-reported severity of difficulty in walking up 10 stairs among participants who were hospitalized at the time of initial illness*

| **Symptom**** | **Any vs. None** OR (95% CI) | **>A Little vs. A Little****** OR (95% CI) | **>Some vs. Some** OR (95% CI) | **Unable vs. A Lot** OR (95% CI) | **Prevalence of Symptom or Factor** | **Proportion of Times Selected by Algorithms***** |
|---|---|---|---|---|---|---|
| Dizziness | **1.55 (1.15, 2.09)** | | | | 0.39 | **1.00** |
| Heavy Limbs | **1.98 (1.38, 2.85)** | | 1.28 (0.78, 2.13) | | 0.21 | 0.67 |
| Sick to Stomach | **1.38 (1.03, 1.85)** | | | | 0.32 | 0.67 |
| Rapid Heart Rate | **1.34 (1.00, 1.79)** | | | | 0.44 | 0.67 |
| Tinnitus | | **3.31 (1.30, 11.2)** | | | 0.10 | 0.67 |
| Skin Problems | | | **2.33 (1.06, 5.48)** | | 0.08 | 0.67 |
| Pneumonia | | | 1.52 (0.88, 2.67) | | 0.21 | 0.67 |
| Disturbed Sleep | | | 1.28 (0.71, 2.32) | | 0.16 | 0.67 |
| Tightness in Chest | | | 1.19 (0.76, 1.86) | | 0.58 | 0.67 |
| Hallucinations | | | **0.32 (0.17, 0.60)** | | 0.10 | 0.67 |
| Pain in Teeth | | | 1.59 (0.88, 2.96) | | 0.13 | 0.33 |
| Fever | | | 1.59 (0.86, 3.04) | | 0.12 | 0.33 |
| Sore Throat | | | 1.35 (0.91, 2.03) | | 0.44 | 0.33 |
| Chills | | | 1.24 (0.80, 1.90) | | 0.58 | 0.33 |

* Models were adjusted for age, gender, BMI, calendar period of infection, activity level prior to infection, and vaccination status
** Three methods for variable selection were used: 1) Random Forest, 2) Multinomial Model with relaxed LASSO, 3) Sequential logistic model with relaxed LASSO; For models with LASSO, the lambda (shrinkage) parameter and gamma (relaxing) parameter were chosen through 20-fold cross validation; Variables included in the model was determined by applying each variable set determined by algorithms and choosing model with the best fit by Akaike's Information Criterion. Final model was sequential logistic model with variable set determined by sequential logistic model with relaxed LASSO
*** Only three persons were boosted with no variability in the outcome and therefore a stable estimate was not obtained
**** Only eight and two persons were fully vaccinated and boosted, respectively, with no variability in outcome and therefore a stable estimate was not obtained

**Table S9.8.** Associations between symptoms at initial COVID-19 illness and self-reported severity of difficulty in doing heavy housework among participants who were not hospitalized at the time of initial illness*

| | **Any vs. None** | **>A Little vs. A Little** | **>Some vs. Some** | **Unable vs. A Lot** | | |
|---|---|---|---|---|---|---|
| **Symptom**** | **OR (95% CI)** | **OR (95% CI)***** | **OR (95% CI)** | **OR (95% CI)** | **Prevalence of Symptom or Factor** | **Proportion of Times Selected by Algorithms**** |
| Heavy Limbs | **1.63 (1.01, 2.72)** | | **2.25 (1.37, 3.80)** | | 0.21 | **1.00** |
| Dizziness | **1.55 (1.07, 2.29)** | | 1.29 (0.86, 1.93) | | 0.39 | **1.00** |
| Sick to Stomach | **1.51 (1.04, 2.23)** | | | | 0.33 | **1.00** |
| Rapid Heart Rate | **1.51 (1.06, 2.18)** | | | | 0.45 | **1.00** |
| Sore Throat | | | 1.33 (0.93, 1.91) | | 0.44 | **1.00** |
| Neuropathy | | | | 1.37 (0.79, 2.36) | 0.12 | **1.00** |
| Fever >100.4 | | | 0.77 (0.54, 1.11) | | 0.53 | **1.00** |
| Difficulty Sleeping | | 0.62 (0.31, 1.22) | | **1.53 (1.04, 2.24)** | 0.37 | **1.00** |
| Lack of Energy | | **2.66 (1.02, 6.83)** | | | 0.87 | 0.67 |
| Tightness in Chest | | 1.78 (0.90,3.51) | | | 0.58 | 0.67 |
| Seizure | | | 3.82 (0.71, 71.3) | | 0.02 | 0.67 |
| Pneumonia | | | 1.36 (0.84, 2.26) | | 0.21 | 0.67 |
| Muscle Ache | | | 1.21 (0.82, 1.80) | | 0.65 | 0.67 |
| Chills | | | 1.09 (0.74, 1.60) | | 0.58 | 0.67 |
| Disturbed Sleep | | | | **1.82 (1.12, 2.94)** | 0.16 | 0.67 |
| Seasonal Allergies | | | 0.60 (0.36, 1.03) | | 0.10 | 0.67 |
| Alopecia | | | **0.50 (0.31, 0.80)** | | 0.21 | 0.67 |
| Altered Smell | | 0.48 (0.19, 1.34) | 1.65 (0.80, 3.68) | | 0.09 | 0.67 |
| Hypersomnia | | 2.10 (0.81, 6.63) | | | 0.13 | 0.33 |
| Cough | | 1.69 (0.90, 3.17) | 0.78 (0.54, 1.13) | | 0.58 | 0.33 |
| Loss of Smell | | 0.67 (0.36, 1.24) | | | 0.46 | 0.33 |
| Headache | | 0.65 (0.30, 1.32) | | | 0.70 | 0.33 |
| Muscle Weakness | | **0.37 (0.14, 0.87)** | | | 0.62 | 0.33 |

* Models were adjusted for age, gender, BMI, calendar period of infection, activity level prior to infection, and vaccination status
** Three methods for variable selection were used: 1) Random Forest, 2) Multinomial Model with relaxed LASSO, 3) Sequential logistic model with relaxed LASSO; For models with LASSO, the lambda (shrinkage) parameter and gamma (relaxing) parameter were chosen through 20-fold cross validation; Variables included in the model was determined by applying each variable set determined by algorithms and choosing model with the best fit by Akaike's Information Criterion. Final model was sequential logistic model with variable set determined by sequential logistic model with relaxed LASSO *** There was no heterogeneity in outcome within one level of exposure of seizure and slow heart rate such that a stable estimate could not be obtained **** There were only six boosted persons without any heterogeneity in outcome and therefore a stable estimate could not be obtained.

**Table S9.9**. Associations between symptoms at initial COVID-19 illness and self-reported severity of difficulty in doing light housework among participants who were hospitalized at the time of initial illness*

| Symptom** | Any vs. None OR (95% CI) | >A Little vs. A Little OR (95% CI) | >Some vs. Some OR (95% CI) | Unable vs. A Lot OR (95% CI) | Prevalence of Symptom or Factor | Proportion of Times Selected by Algorithm** |
|---|---|---|---|---|---|---|
| Heavy Limbs | **1.75 (1.20, 2.58)** | | 1.50 (1.00, 2.27) | | 0.21 | **1.00** |
| Dizziness | **1.58 (1.15, 2.16)** | | | | 0.39 | **1.00** |
| Tremors | 1.37 (0.86, 2.19) | | | | 0.14 | **1.00** |
| Seizure | **3.27 (1.01, 14.7)** | | | | 0.02 | 0.67 |
| Skin Problems | **1.97 (1.07, 3.71)** | | | | 0.08 | 0.67 |
| Tightness in Chest | **1.39 (1.03, 1.88)** | | | | 0.58 | 0.67 |
| Rapid Heart Rate | 1.33 (0.97, 1.81) | | | | 0.45 | 0.67 |
| Sore Throat | 1.21 (0.92, 1.60) | | | | 0.44 | 0.67 |
| Difficulty Sleeping | 1.14 (0.82, 1.58) | | | | 0.37 | 0.67 |
| Hypersomnia | | | **2.27 (1.31, 4.04)** | | 0.13 | 0.67 |
| Fever > 100.4 | **0.71 (0.53, 0.93)** | | | | 0.53 | 0.67 |
| Insomnia | **0.48 (0.30, 0.75)** | | | | 0.15 | 0.67 |
| Disturbed Sleep | 1.42 (0.92, 2.20) | | | | 0.16 | 0.33 |
| Tinnitus | 1.23 (0.74, 2.07) | | | | 0.10 | 0.33 |
| Loss of Taste | **0.75 (0.56, 0.99)** | | | | 0.44 | 0.33 |
| Alopecia | **0.68 (0.46, 1.00)** | | | | 0.21 | 0.33 |

* Models were adjusted for age, gender, BMI, calendar period of infection, activity level prior to infection, and vaccination status
** Three methods for variable selection were used: 1) Random Forest, 2) Multinomial Model with relaxed LASSO, 3) Sequential logistic model with relaxed LASSO; For models with LASSO, the lambda (shrinkage) parameter and gamma (relaxing) parameter were chosen through 20-fold cross validation; Variables included in the model was determined by applying each variable set determined by algorithms and choosing model with the best fit by Akaike's Information Criterion. Final model was sequential logistic model with variable set determined by the sequential logistic regression with relaxed LASSO
*** Only one individual was boosted and therefore a stable estimate was not obtained

**Table S9.10.** Associations between symptoms at initial COVID-19 illness and self-reported severity of mental fatigue as measured by the Woods Mental Fatigue Inventory among participants who were not hospitalized at the time of initial illness*

| Symptom** | Unit Change in WMFI | 95% CI | Prevalence of Symptom or Factor | Proportion of Times Selected By Algorithms** |
|---|---|---|---|---|
| Hallucinations | **2.33** | **(0.22, 4.45)** | 0.10 | **1.00** |
| Tinnitus | 2.14 | (-0.17, 4.44) | 0.10 | **1.00** |
| **Dizziness** | 1.30 | (-0.19, 2.80) | 0.39 | **1.00** |
| Muscle Weakness | 0.70 | (-0.96, 2.36) | 0.62 | **1.00** |
| Joint Aches | 0.51 | (-0.96, 1.98) | 0.50 | **1.00** |
| Tremors | 0.41 | (-1.61, 2.42) | 0.14 | **1.00** |
| Disturbed Sleep | **2.39** | **(0.55, 4.22)** | 0.16 | 0.50 |
| Skin Problems | 2.26 | (-0.33, 4.85) | 0.08 | 0.50 |
| Lack of Energy | **2.10** | **(0.13, 4.08)** | 0.87 | 0.50 |
| Pain in Teeth | 1.61 | (-0.31, 3.53) | 0.13 | 0.50 |
| Tightness in Chest | 0.36 | (-1.08, 1.80) | 0.58 | 0.50 |
| Difficulty Sleeping | 0.26 | (-1.26, 1.77) | 0.37 | 0.50 |

* Models were adjusted for age, gender, BMI, calendar period of infection, activity level prior to infection, and vaccination status

** Two methods for variable selection were used: 1) Random Forest, 2) Gaussian generalized linear model with relaxed LASSO; For model with LASSO, the lambda (shrinkage) parameter and gamma (relaxing) parameter were chosen through 20-fold cross validation; Variables included in the model was determined by applying each variable set determined by algorithms and choosing model with the best fit by Akaike's Information Criterion. Final model was generalized linear model with variable set from the relaxed LASSO

### 10. Physician Diagnoses between Infection and Survey among those with Long-COVID

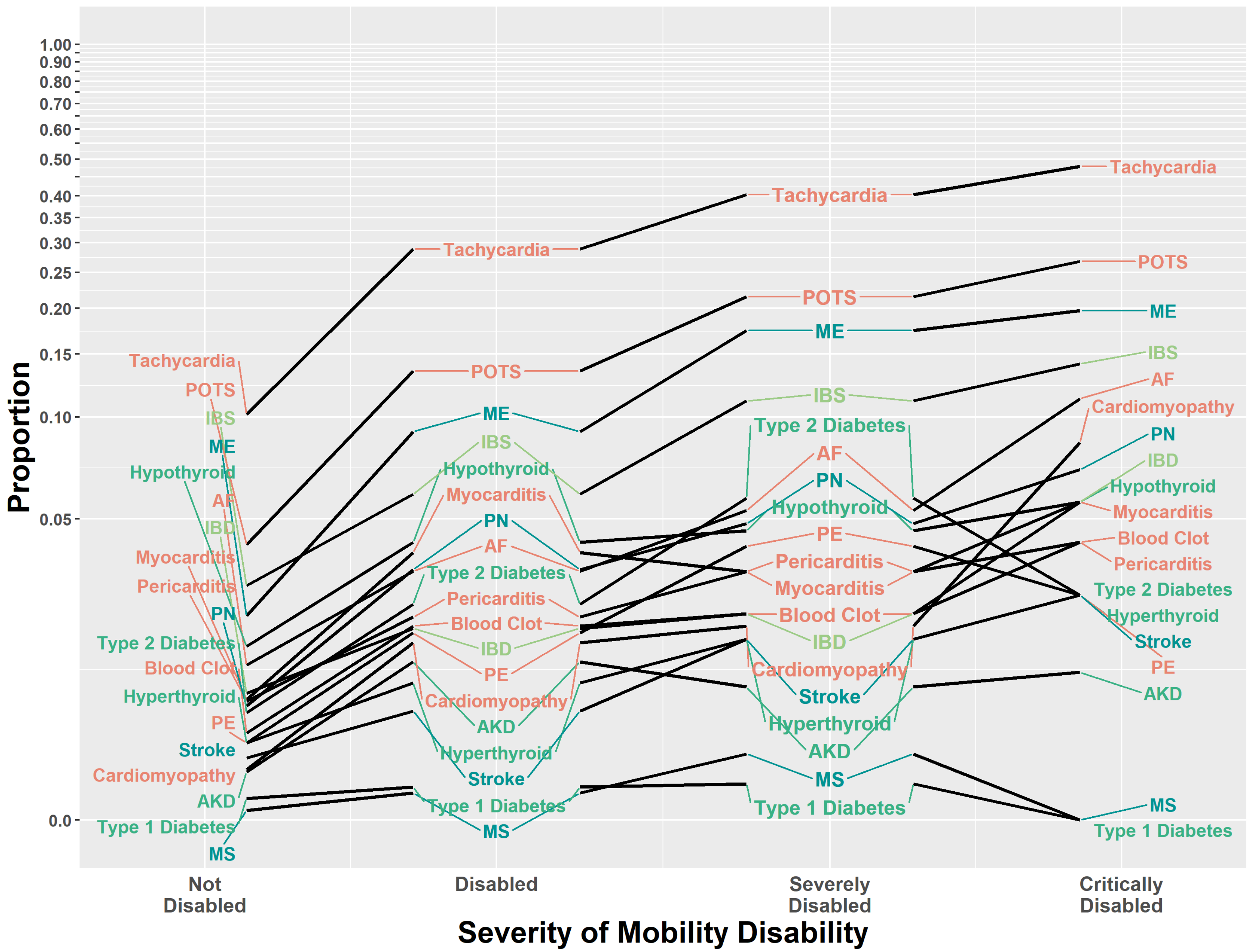

Proportion
1.00
0.90
0.80
0.70
0.60
0.50
0.40
0.35
0.30
0.25
0.20
0.15
0.10
0.05
0.0
Tachycardia
POTS
IBS
ME
Hypothyroid
AF
IBD
Myocarditis
Pericarditis
PN
Type 2 Diabetes
Blood Clot
Hyperthyroid
PE
Stroke
Cardiomyopathy
AKD
Type 1 Diabetes
MS
Not Disabled
Disabled
Severely Disabled
Critically Disabled
Severity of Mobility Disability

**Figure S10.1**. Proportion of individuals diagnosed with conditions by severity of mobility disability. Red are cardiopulmonary diagnoses (AF - atrial fibrillation, Blood Clot, Cardiomyopathy, Pericarditis, PE – pulmonary embolism, POTS – postural orthostatic tachycardia syndrome, Myocarditis, Tachycardia), light green are gastrointestinal (Irritable Bowel Disease, Irritable Bowel Syndrome), blue-green are neurologic diagnoses (MS – multiple sclerosis, ME – myaligic encephalomyelitis/chronic fatigue syndrome, PN – peripheral neuropathy, Stroke), and dark green are metabolic/renal diagnoses (AKD - acute kidney disease, Hyperthyroid, Hypothyroid, Type 1 Diabetes, Type 2 Diabetes).

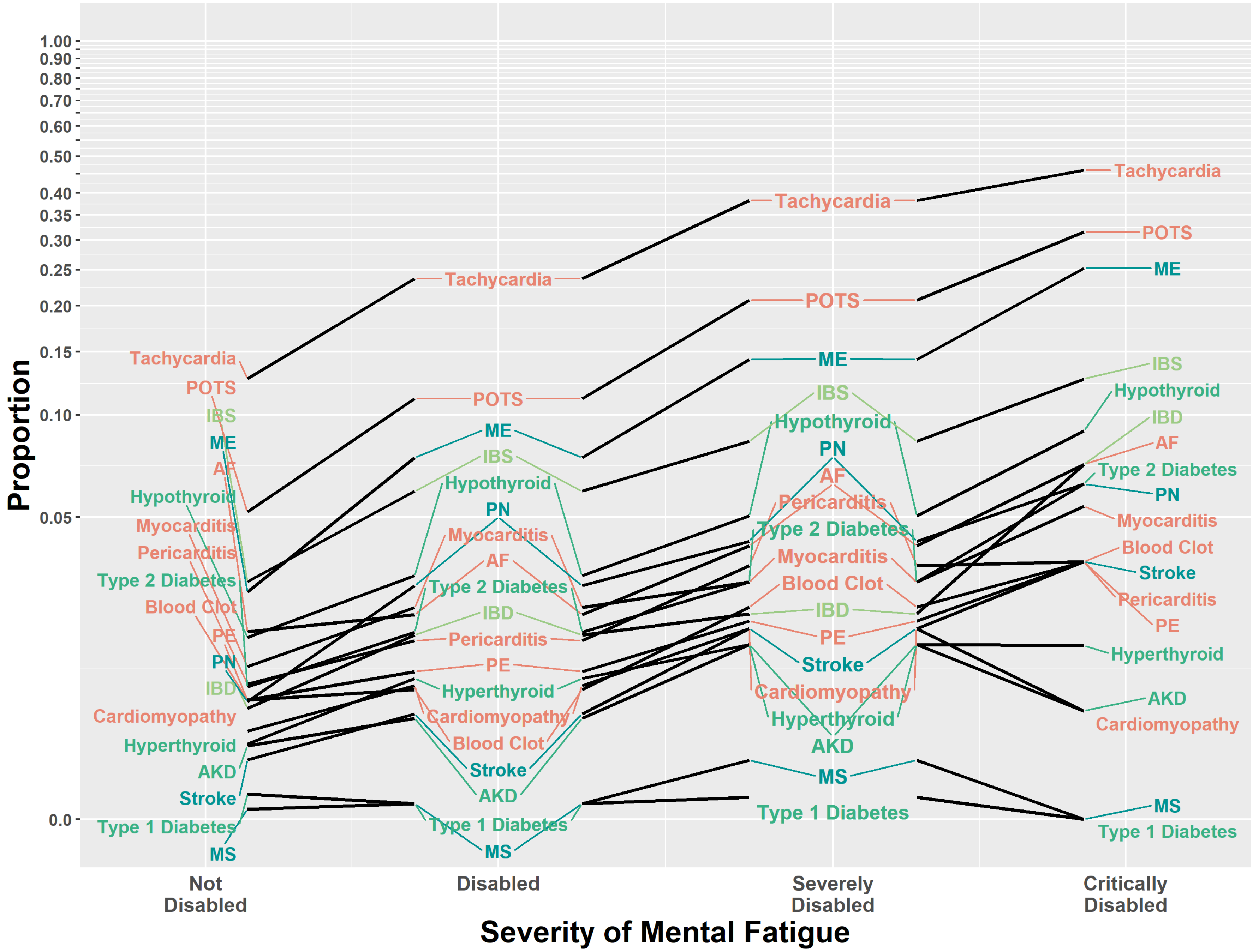

Proportion
1.00
0.90
0.80
0.70
0.60
0.50
0.40
0.35
0.30
0.25
0.20
0.15
0.10
0.05
0.0
Not Disabled
Disabled
Severely Disabled
Critically Disabled
Severity of Mental Fatigue
Tachycardia
POTS
IBS
ME
AF
Hypothyroid
Myocarditis
Pericarditis
Type 2 Diabetes
Blood Clot
PE
PN
IBD
Cardiomyopathy
Hyperthyroid
AKD
Stroke
Type 1 Diabetes
MS

**Figure S10.3**. Proportion of individuals diagnosed with conditions by severity of mental fatigue. Red are cardiopulmonary diagnoses (AF - atrial fibrillation, Blood Clot, Cardiomyopathy, Pericarditis, PE – pulmonary embolism, POTS – postural orthostatic tachycardia syndrome, Myocarditis, Tachycardia), light green are gastrointestinal (Irritable Bowel Disease, Irritable Bowel Syndrome), blue-green are neurologic diagnoses (MS – multiple sclerosis, ME – myaligic encephalomyelitis/chronic fatigue syndrome, PN – peripheral neuropathy, Stroke), and dark green are metabolic/renal diagnoses (AKD - acute kidney disease, Hyperthyroid, Hypothyroid, Type 1 Diabetes, Type 2 Diabetes).
